# Long-read RNA-seq atlas of novel microglia isoforms elucidates disease-associated genetic regulation of splicing

**DOI:** 10.1101/2023.12.01.23299073

**Authors:** Jack Humphrey, Erica Brophy, Roman Kosoy, Biao Zeng, Elena Coccia, Daniele Mattei, Ashvin Ravi, Anastasia G. Efthymiou, Elisa Navarro, Benjamin Z. Muller, Gijsje JLJ Snijders, Amanda Allan, Alexandra Münch, Reta Birhanu Kitata, Steven P Kleopoulos, Stathis Argyriou, Zhiping Shao, Nancy Francoeur, Chia-Feng Tsai, Marina A Gritsenko, Matthew E Monroe, Vanessa L Paurus, Karl K Weitz, Tujin Shi, Robert Sebra, Tao Liu, Lot D. de Witte, Alison M. Goate, David A. Bennett, Vahram Haroutunian, Gabriel E. Hoffman, John F. Fullard, Panos Roussos, Towfique Raj

**Affiliations:** Department of Genetics and Genomic Sciences, Icahn School of Medicine at Mount Sinai, New York, NY, USA; Nash Family Department of Neuroscience & Friedman Brain Institute, Icahn School of Medicine at Mount Sinai, New York, NY, USA; Ronald M. Loeb Center for Alzheimer’s Disease, Icahn School of Medicine at Mount Sinai, New York, NY, USA; Icahn Genomics Institute, Icahn School of Medicine at Mount Sinai, New York, NY, USA; Estelle and Daniel Maggin Department of Neurology, Icahn School of Medicine at Mount Sinai, New York, NY, USA; Center for Disease Neurogenomics, Icahn School of Medicine at Mount Sinai, New York, USA; Department of Psychiatry, Icahn School of Medicine at Mount Sinai, New York, USA; Department of Biochemistry and Molecular Biology, Faculty of Medicine (Universidad Complutense de Madrid), Madrid, Spain; Centro de Investigación Biomédica en Red sobre Enfermedades Neurodegenerativas (CIBERNED), Madrid, Spain; Instituto Ramon y Cajal de Investigacion Sanitaria (IRYCIS), Madrid, Spain; Biological Sciences Division, Pacific Northwest National Laboratory, Richland, Washington, USA; Black Family Stem Cell Institute, Icahn School of Medicine at Mount Sinai, New York, NY, 10029, USA; Global Health and Emerging Pathogens Institute, Icahn School of Medicine at Mount Sinai, New York, NY, 10029, USA; Rush Alzheimer’s Disease Center, Rush University Medical Center, Chicago, Illinois, USA; Mental Illness Research Education, and Clinical Center (VISN 2 South), James J. Peters VA Medical Center, Bronx, NY, USA

## Abstract

Microglia, the innate immune cells of the central nervous system, have been genetically implicated in multiple neurodegenerative diseases. We previously mapped the genetic regulation of gene expression and mRNA splicing in human microglia, identifying several loci where common genetic variants in microglia-specific regulatory elements explain disease risk loci identified by GWAS. However, identifying genetic effects on splicing has been challenging due to the use of short sequencing reads to identify causal isoforms. Here we present the isoform-centric microglia genomic atlas (isoMiGA) which leverages the power of long-read RNA-seq to identify 35,879 novel microglia isoforms. We show that the novel microglia isoforms are involved in stimulation response and brain region specificity. We then quantified the expression of both known and novel isoforms in a multi-ethnic meta-analysis of 555 human microglia short-read RNA-seq samples from 391 donors, the largest to date, and found associations with genetic risk loci in Alzheimer’s disease and Parkinson’s disease. We nominate several loci that may act through complex changes in isoform and splice site usage.

## Introduction

Microglia are the myeloid innate immune cells of the central nervous system. The past decade has seen increasing genetic evidence, from both rare variants found in sequencing studies and common variants found in genome-wide association studies (GWAS), implicating the myeloid cell lineage in the pathogenesis of multiple neurodegenerative diseases, chief among them Alzheimer’s disease (AD) and Parkinson’s disease (PD) (Guerreiro et al. 2013; Lambert et al. 2013; Towfique Raj et al. 2014; T. Raj et al. 2014; Efthymiou and Goate 2017; Huang et al. 2017; Schwartzentruber et al. 2021; Andersen et al. 2021; Novikova et al. 2021; Navarro et al. 2021). Furthermore, recent studies have shown that several genetic loci can be colocalized to epigenetic annotations in microglia (Nott et al. 2019; Corces et al. 2020; Young et al. 2021; Lopes et al. 2022; Kosoy et al. 2022; Bryois et al. 2022; Fujita et al. 2022; Schilder and Raj 2022; Langston et al. 2022). Therefore, backed up by robust genetic associations, microglia are a key cell-type of interest for the identification of new therapeutic targets in AD and PD.

Previous studies of microglia have combined genetic and transcriptomic data to find associations between common genetic variants and gene expression: namely expression quantitative trait loci (eQTLs). By integrating GWAS data with eQTLs and epigenetic annotations, multiple risk loci have been found to influence microglia gene expression through modification of microglia-specific enhancer sequences (Young et al. 2021; Lopes et al. 2022; Kosoy et al. 2022; Nott et al. 2019; Corces et al. 2020). However, the individual unit of the transcriptional program is the mRNA transcript or isoform, which results from a combination of molecular decisions in promoter usage, alternative 5’/3’ splice site usage, exon usage, intron retention and polyadenylation site, all of which can be modified by *cis*-acting DNA variants. Genetic associations with splicing can be identified by the mapping of splicing quantitative trait loci (sQTLs). As opposed to directly affecting gene expression, multiple GWAS loci have been associated with intron splicing, promoter choice, and polyadenylation (Y. I. Li et al. 2016; GTEx Consortium 2020; Mittleman et al. 2020; Alasoo et al. 2019; Y. I. Li et al. 2018; Towfique Raj et al. 2018).

The current state of the art for sQTL mapping, Leafcutter, relies on deriving ratios of overlapping intron-junction-spanning reads (Y. I. Li et al. 2018). This approach is flexible and easily scalable to 1000s of samples, but does not pinpoint the specific isoforms involved, and does not include intronic or exonic read coverage, necessary for finding genetic associations with intron retention or polyadenylation site choice. We and others previously used Leafcutter to identify sQTLs in *CD33* (Young et al. 2021; Lopes et al. 2022) and *MS4A6A (Lopes et al. 2022)* in human microglia colocalizing with AD GWAS, and *FAM49B* (now known as *CYRIB*) in human monocytes colocalizing with PD risk (Navarro et al. 2021). Alternative sQTL mapping approaches, which rely on estimating isoform expression (Monlong et al. 2014; Qi et al. 2022), or grouping isoforms into sets of splicing events (Alasoo et al. 2019; Y. Zhang et al. 2020), have identified additional sources of genetic regulation. However, these methods suffer by being limited only to isoforms that have already been discovered in references such as GENCODE, and therefore may miss rarer isoforms expressed in specific contexts or cell-types that are currently unannotated (D. Zhang et al. 2020).

Long-read RNA-seq facilitates the capture of entire mRNA molecules in a single sequencing read, promising the ability to identify novel isoforms and open reading frames (ORFs) (Sharon et al. 2013; Castaldi et al. 2022; Leung et al. 2021). Although it is now feasible to map genetic associations purely using long-read RNA-seq samples (Glinos et al. 2022), a more cost-effective approach is to combine long-read-derived isoform information with the high sequencing depth and large cohort sizes more commonly found with short-read RNA-seq (Abood et al. 2023). Here, we extend our previous microglia QTL studies (Lopes et al. 2022; Kosoy et al. 2022) by generating long-read RNA-seq to identify novel isoforms in order to better interpret the genetic regulation of splicing in microglia. We present the isoform-centric microglia genomic atlas (isoMiGA), generated using long reads from postmortem microglia samples to identify novel isoforms. isoMiGA is able to augment the standard GENCODE transcriptome reference to quantify their expression in both existing and newly generated short-read microglia RNA-seq samples. This provides an increased ability to both discover and interpret the genetic regulation of mRNA splicing in microglia as well as its links to neurodegenerative disease.

## Results

### Long-read RNA sequencing of human microglia identifies novel isoforms

We generated long-read RNA-seq data using the IsoSeq (Pacific Biosciences) Sequel II instrument and circular consensus sequencing (CCS) reads, sequencing 30 single-molecule real time (SMRT) 8M SMRTcells using bulk mRNA from microglia purified from postmortem human brains, the majority from prefrontal cortex and medial frontal gyrus tissues, combining multiple diagnoses, including AD and Lewy body dementia, as well as non-neurological control samples (**Figure 1; Supplementary Fig. 1; Supplementary Table** generated a total of 89,285,858 full-length long reads, of which 99.73% uniquely aligned to the **1**). We human reference genome (hg38). Median read lengths ranged from 558-3792 bases, with 28 of 30 samples having a median CCS read length greater than 2 kbp (**Supplementary Fig. 2**). We then performed hybrid assembly using Stringtie2 (Shumate et al. 2022; Kovaka et al. 2019), pairing each long-read sample with its corresponding short-read sample generated using conventional Illumina RNA-seq. After quality control (**Supplementary Fig. 3**), removing potentially artifactual isoforms due to internal polyA priming (Roy and Chanfreau 2020) or reverse-transcription (Verwilt, Mestdagh, and Vandesompele 2023), we were able to assemble 128,436 isoforms in 25,956 genes, of which 35,879 isoforms and 2,238 genes were novel (not found in GENCODE v38), whereas 92,557 exactly matched an annotated transcript (**Figure 2a**). Collapsing the isoforms into splicing events, we observed that alternate promoter usage was the most common splicing event, and that compared to the GENCODE reference, intron retention events were highly abundant (**Figure 2b**). Classifying novel isoforms by their relationship with annotated isoforms using the SQANTI (v3) (Tardaguila et al. 2018) framework (**Figure 2a; Supplementary Table 2**), we found that, while 92,557 isoforms were full splice matches (FSM) to annotated isoforms, 32,459 of the novel isoforms contained partial sequence matches to known isoforms. This included 18,727 novel-in-catalog isoforms, where a novel transcript uses a novel arrangement of annotated splice sites, 11,607 novel-not-in-catalog isoforms, where a novel transcript includes novel splice sites not found in any annotated transcript, and 2,142 incomplete splice matches, where the novel isoform matches a 5’ or 3’ fragment of a known transcript. We observed 1,725 readthrough fusion isoforms, containing sequences spliced together linearly from 2, or up to 6, adjacent genes. Of the 590 readthrough fusion genes, 36 were previously identified in the Conjoined Genes database (Prakash et al. 2010). Additionally, we observed 731 intergenic isoforms, 621 antisense isoforms, containing sequences transcribed antisense to a known gene, and 326 genic isoforms found within introns of known genes.

**Fig. 1.**
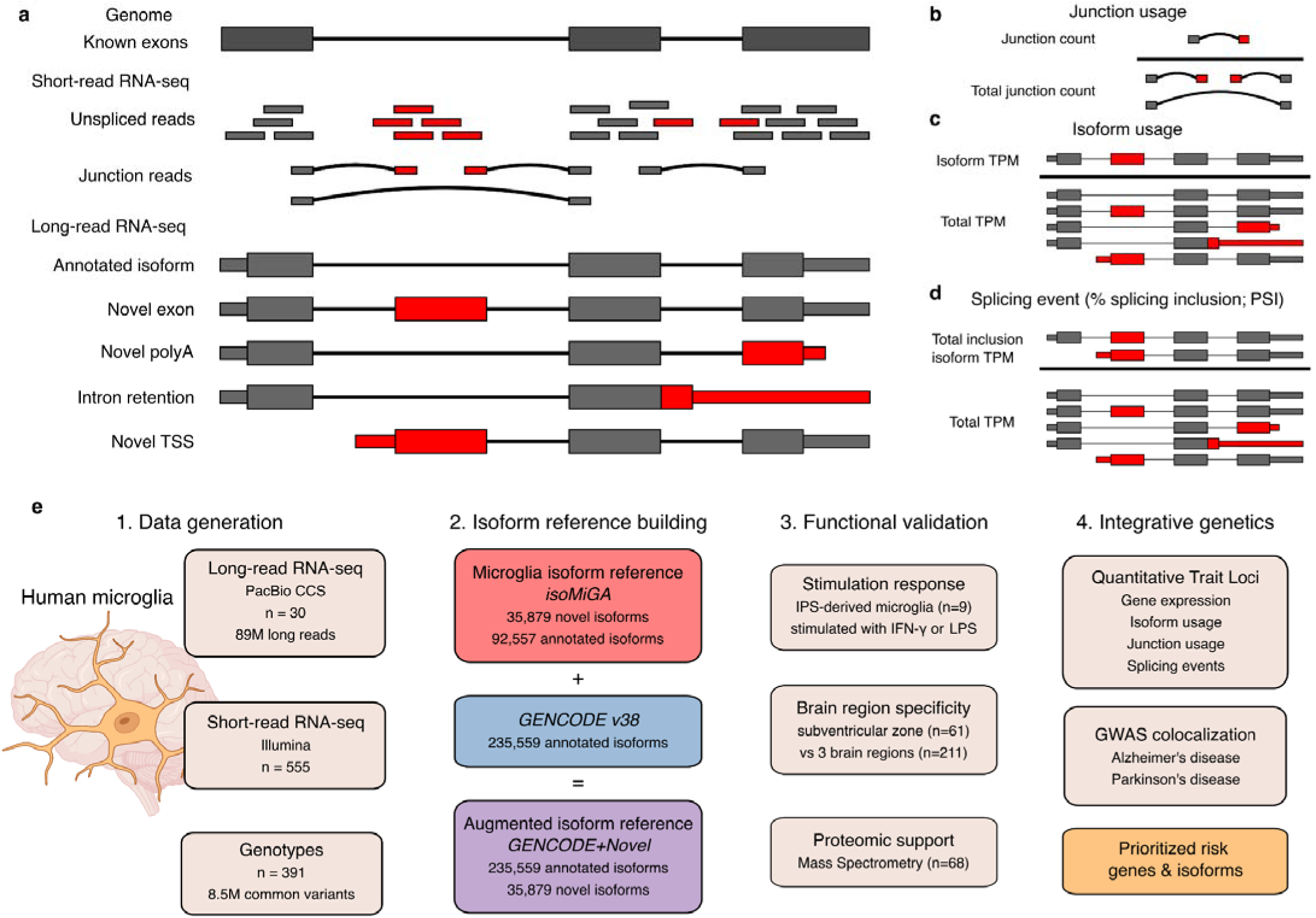
The isoform-centric microglia genomic atlas (isoMiGA) project. **a,** Comparison of short-read and long-read RNA-seq for novel isoform discovery. Short-read RNA-seq relies on spliced junction reads to define splice sites, whereas long-read RNA-seq can sequence an entire isoform in a single read, enabling the precise delineation of isoform structure, identifying novel sequences not present in the reference annotation (red boxes) as well as the potential amino acid coding sequence for each isoform, represented by the wide boxes. Narrow boxes represent 5’ and 3’ untranslated regions. **b**, Junction usage, as exemplified by Leafcutter, takes a ratio of each junction over the total set of overlapping junctions. **c**, Isoform usage calculates the ratio of each isoform over the total set of isoforms. **d,** In splicing events, as exemplified by SUPPA, the isoforms that contain the inclusion of a particular splicing event (in this case, inclusion of novel cassette exon), are summed and divided by the total isoforms for that gene. **e**, Schematic outline of data generation and analysis in this study. TSS: transcription start site, polyA: polyadenylation site.

**Fig. 2.**
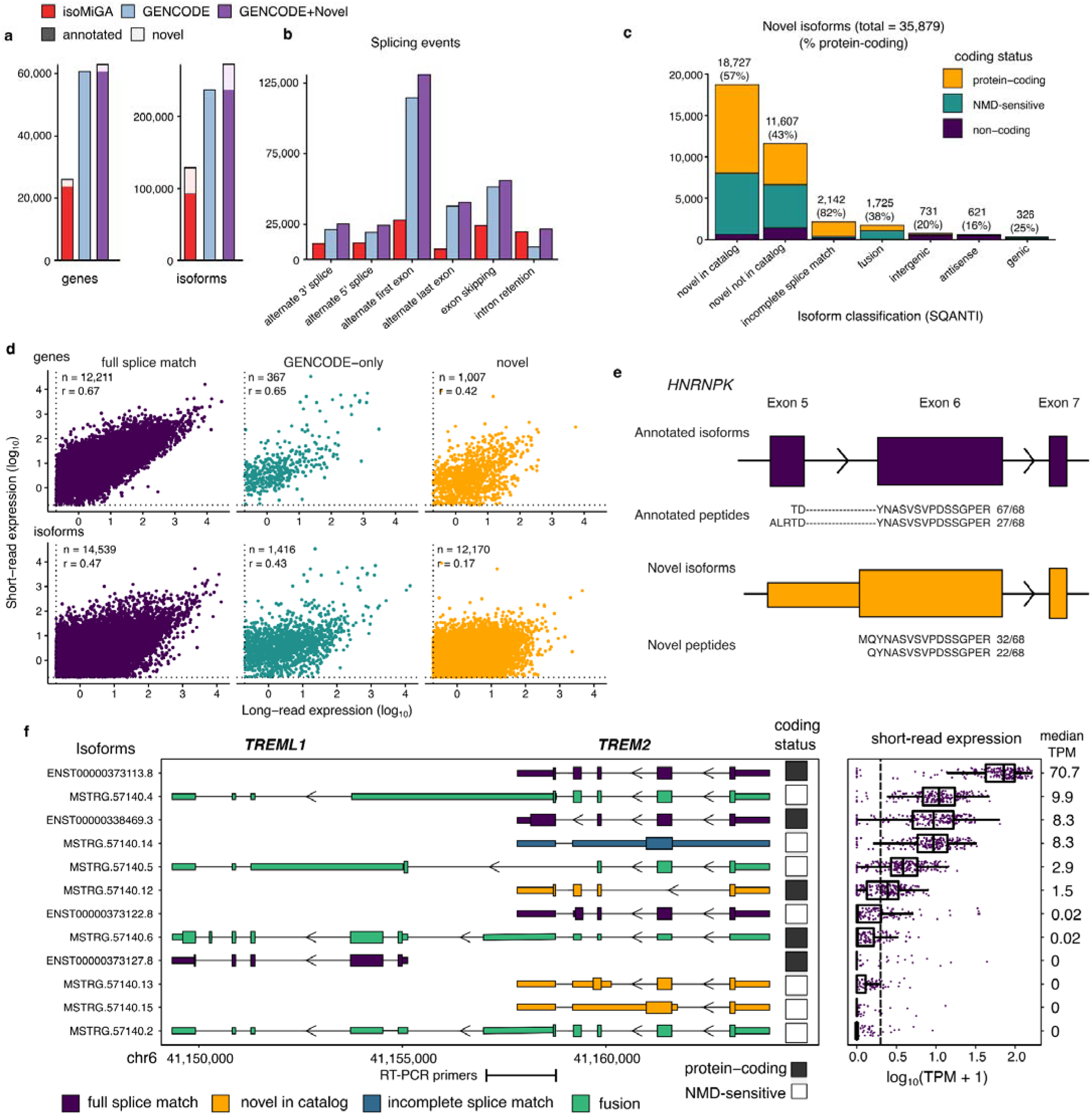
Identifying novel isoforms in human microglia. **a,** Comparing numbers of genes and isoforms found in the microglia long-read samples (isoMiGA), with GENCODE, and an augmented reference combining GENCODE annotated isoforms with the novel isoforms found in isoform (GENCODE+Novel). **b,** Numbers of splicing events identified in the three references by SUPPA. **c**, Distribution and inferred properties of each class of novel isoform, grouped by coding status. Number of each isoform type plotted, with the percentage of isoforms predicted to be protein-coding in parentheses. NMD: nonsense-mediated decay. **d,** Correlations between long-read and short-read expression of genes (upper panels) and isoforms (lower panels), split by whether annotated isoform seen in long-read (annotated), only in GENCODE (GENCODE-only) or only in long-read reference (novel). Expression summarized as median TPM in long-read samples (n=30) and largest short-read microglia cohort (n=185). n refers to the number of genes or isoforms with median TPM > 0.1 in both sequencing modalities, r is the Pearson’s correlation coefficient. **e**, Shotgun proteomics-derived peptides support a novel downstream translation start site in exon 6 of *HNRNPK*. Numbers refer to the number of mass spectrometry samples the peptide was detected in. **f,** Isoforms discovered in the *TREM2/TREML1* locus include multiple novel isoforms, including fusion isoforms connecting the two genes, most of which are not predicted to be translated (NMD-sensitive). The structure of each isoform is shown with wider boxes denoting the predicted coding sequence and narrower boxes depicting non-coding sequence. All isoforms are transcribed in the negative direction, denoted by the arrows. Introns shortened to better display exon structure. Locations of RT-PCR primers confirming gene fusion located at bottom of plot. **d**, The expression of each isoform in short-read RNA-seq microglia (n=185). Boxplots plot the first quartile, the median and the third quartile of the values, with the whiskers denoting 1.5 times the interquartile range. Overlaid violins plot the range and distribution of the values.

We assessed each isoform for potential protein-coding ability using GeneMarks (v5.1) to identify ORFs (Lukashin and Borodovsky 1998). Additionally, any isoform with a stop codon following a splice site was predicted by SQANTI to be degraded by nonsense-mediated decay (NMD). We observed that incomplete splice match isoforms had the highest proportion of predicted protein-coding isoforms (82%) (**Figure 2c**; **Table 1**), followed by novel in catalog isoforms (57%). The majority of the other transcript types were predicted to be non-coding or sensitive to NMD. We overlapped the transcript start coordinates of our isoforms with ATAC-seq data from postmortem human microglia (Kosoy et al. 2022) (**Table 1**) to evaluate whether the TSS positions overlapped putative promoter sequences. Multiple novel isoform types had a greater rate of TSS over ap than the annotated FSM isoforms.

**Table 1:** All isoforms discovered in microglia. NMD: nonsense-mediated decay; TSS: transcription start site. ATAC: assay for transposase-accessible chromatin.

| Isoform type | number | protein-coding | NMD-sensitive | TSS overlaps microglia ATAC-seq (Kosoy et al. 2022) |
| --- | --- | --- | --- | --- |
| Full splice match | 92,557 | 63% | 12% | 57% |
| Incomplete splice match | 2,142 | 82% | 9.2% | 74% |
| Novel in catalog | 18,727 | 57% | 39% | 82% |
| Novel not in catalog | 11,607 | 43% | 45% | 70% |
| Fusion | 1,725 | 38% | 58% | 67% |
| Intergenic | 731 | 20% | 12% | 18% |
| Antisense | 621 | 16% | 12% | 10% |
| Genic | 326 | 25% | 38% | 61% |

We reasoned that we may be missing isoforms that are expressed in microglia but not captured in the long-read RNA-seq reference, due to comparatively lower read depth per sample in the long-read RNA-seq compared to standard short-read RNA-seq, or to differences in gene capture, as long-read RNA-seq requires enrichment of polyadenylated mRNA through oligo dT capture, whereas short-read RNA-seq employs either oligo dT capture or total RNA capture followed by ribosomal RNA depletion. We therefore combined our 35,879 novel isoforms with the full set of 235,559 annotated isoforms found in the GENCODE v38 release, and quantified the expression of each isoform in all 30 long-read RNA-seq samples, along with a set of 555 purified microglia short-read RNA-seq samples comprised from three published studies (Young et al. 2021; Lopes et al. 2022; Kosoy et al. 2022) as well as an additional 121 samples newly generated for this study (**Supplementary Table 3**). For each cohort, we calculated the median isoform and gene expression in transcripts per million (TPM), observing that a minority of isoforms (1,416) found in GENCODE, but not in the long-read reference (“GENCODE-only”), were quantifiable in both long and short-read RNA-seq at a median TPM > 0.1. Whereas 93% of genes found in the long-read reference were either protein-coding genes or lncRNAs, 43% of GENCODE-only genes were non-polyadenylated RNA species such as microRNAs and small nucleolar RNAs (**Supplementary** Fig 4). Comparing the median TPMs for genes and isoforms between short and long-read RNA-seq cohorts, we found them to be highly correlated, with annotated isoforms found in both long-read and in GENCODE (“full splice match”) having the highest correlation (**Figure 2d**). We next compared the expression of each isoform for the largest individual short-read RNA-seq cohort (n=185; ribo-depleted total RNA) (**Supplementary** Fig 4). Comparing the 92,557 annotated isoforms that we detected in the long-read RNA-seq assembly to the 143,201 annotated isoforms found only in GENCODE, we observed that only a minority (14%) of the undetected GENCODE-only isoforms were expressed in the short-read RNA-seq samples at the common threshold of TPM > 1. This suggests that, while long-read RNA-seq did not capture every transcript and gene expressed in microglia, it captured the vast majority. Surprisingly, we observed that the novel isoforms as a group are more highly expressed than the annotated isoforms, with only the antisense isoforms having a lower expression.

To confirm whether any of our potentially protein-coding isoforms produced functional proteins, we generated shotgun mass spectrometry proteomics data on 68 unique human microglia samples, producing 137,704 unique trypsin-digested peptides (**Supplementary Table 4;** *Mattei et al, in preparation*). By matching the peptide fragments to our predicted coding sequences, 69,540 peptides matched only to annotated isoforms and 68,095 to sequences shared by both annotated and novel isoforms. 69 peptides matched exclusively to 112 novel isoforms within 65 genes. The novel peptides as a set were less abundant and were found in fewer spectrometry runs (**Supplementary Fig. 5**). Looking at genes with at least two novel peptide matches, we observed several RNA-binding protein genes, including *HNRNPK, HNRNPH1, SRRM1*, and *FXR2*. We highlight a set of novel isoforms in *HNRNPK*, an RNA-binding protein recently associated with the aging brain (Bampton et al. 2021), which contains a novel intron retention event between exons 5 and 6, leading to a novel start codon upstream of the canonical exon 6 coding domain. We found two peptides supporting the canonical exon 5-6 splicing and two novel peptides supporting the novel upstream start site (**Figure 2e**).

To demonstrate the diversity of isoforms discovered in the isoMiGA reference, we present the isoforms found in *TREM2* (**Figure 2c**) and *CD33* (**Supplementary Fig. 6**), two genes with robust genetic associations to AD (Guerreiro et al. 2013; T. Raj et al. 2014). Strikingly, both loci contain detectable readthrough fusion isoforms that connect two genes together. The *TREM2*/*TREML1* region has 12 detectable isoforms in IsoMiGA, 4 of which are found in GENCODE, and 8 of which have a median short-read expression > 0 TPM (**Figure 2f**), although the major isoform (median short-read TPM=70) is the canonical *TREM2* transcript. 4 of the novel isoforms (MSTRG.54710.2, MSTRG.54710.4, MSTRG.54710.5, MSTRG.54710.6) splice from the final exon of *TREM2* into the region containing *TREML1.* We validated this fusion by reverse transcription-PCR (RT-PCR) using primers that span the extension of the *TREM2* 3’UTR (**Supplementary Fig. 7**). Although 3 of the fusion isoforms are predicted to be non-coding through NMD, including the highest expressed fusion MSTRG.54710.4, one isoform MSTRG.54710.6 is predicted to contain a coding ORF containing 6 exons of *TREML1*, with the *TREM2* exons acting as a 5’UTR. This isoform is more highly expressed in short-read RNA-seq than the canonical *TREML1* isoform itself. Another novel isoform, MSTRG.54710.12, involves skipping of exon 2 which encodes the ligand-binding domain, has recently been reported by two other groups (Kiianitsa et al. 2021; Shaw et al. 2022). In *CD33*, as well as observing the known exon 2 skipping isoforms, we also observed multiple novel isoforms (**Supplementary** Fig 6), including two predicted coding isoforms retaining the intron between exon 1 and 2 (MSTRG.34112.20, MSTRG.34112.16) previously observed in leukocytes (Malik et al. 2015). We predicted this would create a novel downstream ORF, as well as multiple readthrough fusion isoforms splicing exons from the upstream pseudogene *SIGLEC22P* into *CD33* (MSTRG.34112.2 etc.), creating a novel 5’UTR, which we validated with RT-PCR (**Supplementary** Fig 7).

### Novel isoforms have stimulation, region and subtype specificity

Microglia occupy a variety of different states within the brain, responding to changes in their micro-environment, including inflammatory stimuli. If any of our novel microglia isoforms play a functional role in microglia, we would expect to see variation in their expression in response to external stimuli and between different regions of the brain (Grabert et al. 2016; van der Poel et al. 2019). We generated induced pluripotent stem cell-derived microglia (iMGLs) from 3 isogenic lines harboring a Parkinson’s disease linked mutation (G2019S) in the *LRRK2* gene generated from a single human donor and treated them with either lipopolysaccharide (LPS) or interferon gamma (IFN-γ) for 24 hours, to simulate a reaction to bacterial or viral infection, respectively (**Figure 3a**) (*Navarro et al., in preparation)*. We estimated isoform expression in short-read RNA-seq data from three independent differentiations of these samples to an augmented reference containing both GENCODE and our novel isoforms, with 17,522 annotated and 872 novel isoforms expressed with a median TPM > 0.1. Novel genes in our reference come in 3 categories: antisense, gene fusions, and fully novel genes (isoforms not associated with any annotated gene). We performed differential expression, identifying 2,192 annotated and 75 novel genes to be differentially expressed by IFN-γ treatment (|LFC| > 1; FDR < 0.05) (**Figure 3b; Supplementary Table 5**), whereas 539 annotated and 28 novel genes were differentially expressed by LPS treatment (**Supplementary Fig. 8; Supplementary Table 6**). Of the novel genes, 18 were observed in both IFN-γ and LPS treatment, including the *TREML1+TREM2* readthrough fusion isoform, which was downregulated by both stimuli with a stronger effect size than the *TREM2* gene itself. Testing differential isoform usage (DIU) compares the relative expression of each isoform to the total expression of each gene, and allows for differences in specific isoforms to be examined. We tested 13,187 annotated and 4,589 novel isoforms for DIU, and only observed significant isoforms in response to IFN-γ, with 42 annotated isoforms and 9 novel isoforms shifting usage (FDR < 0.05) (**Figure 3c; Supplementary Table 7)**. These results suggest that some of the novel isoforms identified play a role in inflammatory response and may have specific functions.

**Fig. 3.**
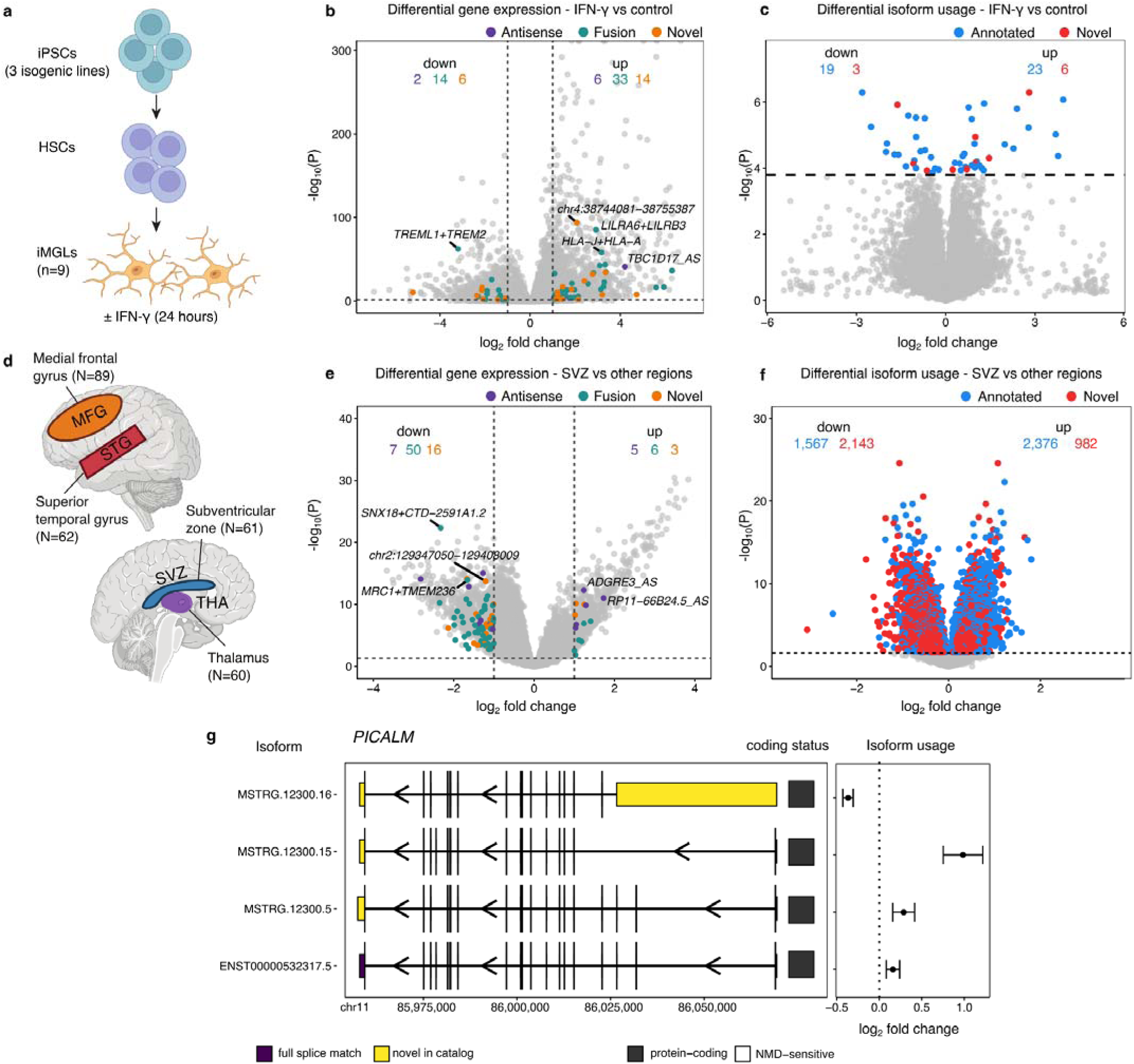
Differential gene expression and isoform usage in interferon stimulation and between brain regions finds novel genes and isoforms. **a**, Schematic for generation of iPS-derived microglia and stimulation procedures. **b,** Differential gene expression finds multiple types of novel genes altered. **c,** Differential isoform usage finds novel isoforms altered. **d**, Schematic of the comparison between microglia from four brain regions. **e**, Differential expression finds novel genes mostly downregulated in microglia from the SVZ. **f,** Differential isoform usage finds novel isoforms downregulated compared to annotated isoforms in SVZ microglia. **g**, The AD-associated gene *PICALM* has region-specific differential isoform usage of 4 isoforms.

Our previous study comparing microglia purified from four different brain regions found widespread region-specific differential gene expression and transcript usage between microglia from cortical and non-cortical regions of the human brain (**Figure 3d**), with microglia from the subventricular zone (SVZ) being particularly distinct (Lopes et al. 2022). We repeated this analysis with our novel isoforms, comparing 61 SVZ microglia to 211 samples from the medial frontal gyrus, superior temporal gyrus and the thalamus. Of the 17,522 annotated and 872 genes tested, we observed 1,217 annotated and 87 novel genes to be differentially expressed (|LFC| > 1; FDR < 0.05) (**Figure 3e; Supplementary Table 8**). DTU analysis identified 3,943 annotated and 3,125 novel isoforms differentially used by SVZ microglia (FDR < 0.05), with novel isoforms being preferentially downregulated in the SVZ (**Figure 3f; Supplementary Table 9**). This included the AD risk gene *PICALM*, where 4 isoforms were differentially used, with the novel protein-coding isoform MSTRG.12300.16, which lacks the upstream 3 coding exons, being downregulated in SVZ microglia relative to the rest of the isoforms (**Figure 3g**).

### Novel isoforms enhance splicing QTL discovery

Combining multiple matched genotype and transcriptomic cohorts increases discovery of cis-genetic regulatory associations (Kosoy et al. 2022). We combined our short-read microglia data for a cis-quantitative tr it locus (QTL) analysis of 555 short-read RNA-seq samples from 391 donors, forming the largest microglia paired genetic/transcriptomic cohort to date. As 64 of the donors were of non-European ancestry (**Supplementary** Fig 9), and our cohorts included repeated donors across multiple brain regions, we applied the multi-ancestry mixed model QTL meta-analysis tool mmQTL (B. Zeng et al. 2022) to include donors and samples of all ancestries in a single random-effects meta-analysis to maximize our discovery power.

We mapped cis-genetic associations with common genetic variants (minor allele frequency > 1%) in a 1 Mbp window with both gene expression (eQTLs) and a range of splicing phenotypes (sQTLs) using both the standard GENCODE transcript annotation and a combined transcript reference including our additional 35,879 novel isoforms (GENCODE+Novel). For studying the genetics of splicing, we mapped the standard junction usage splicing QTLs (juQTLs) using Leafcutter (Y. I. Li et al. 2018). We used the estimated transcript expression in each cohort to map transcript usage QTLs, the relative expression of each transcript compared to the total expression for each gene (tuQTLs). Finally, we used SUPPA (Trincado et al. 2018) to decompose the transcript references into sets of splicing events, including cassette exons (SE), intron retention (RI), alternate start (AF) and end sites (AL), and alternate 5’ (A5) and 3’ splice sites (A3). This combined approach allowed us to capture the genetic regulation of mRNA at multiple levels.

We mapped each QTL type using either the standard GENCODE reference or the augmented GENCODE+Novel reference. Increasing the number of testable isoforms increases the number of tests performed, as well as alters minimum TPM expression thresholds. Comparing QTL discovery rates at qvalue < 0.05 between the two transcript references, we observed an increase in QTL discovery of transcript usage and splicing events, but a reduction in eQTL discovery rate with the augmented GENCODE+Novel reference. (**Figure 4a**). Genes with eQTLs only found in the GENCODE reference did not differ in the total number of annotated or known isoforms compared to eQTLs found only in the GENCODE+Novel reference and were highly concordant in their effect directions between references (**Supplementary Fig. 10**). Despite this, we identified 456 novel genes to have an eQTL and 5,658 novel isoforms in 3,545 genes to have a tuQTL at qvalue < 0.05.

**Fig. 4.**
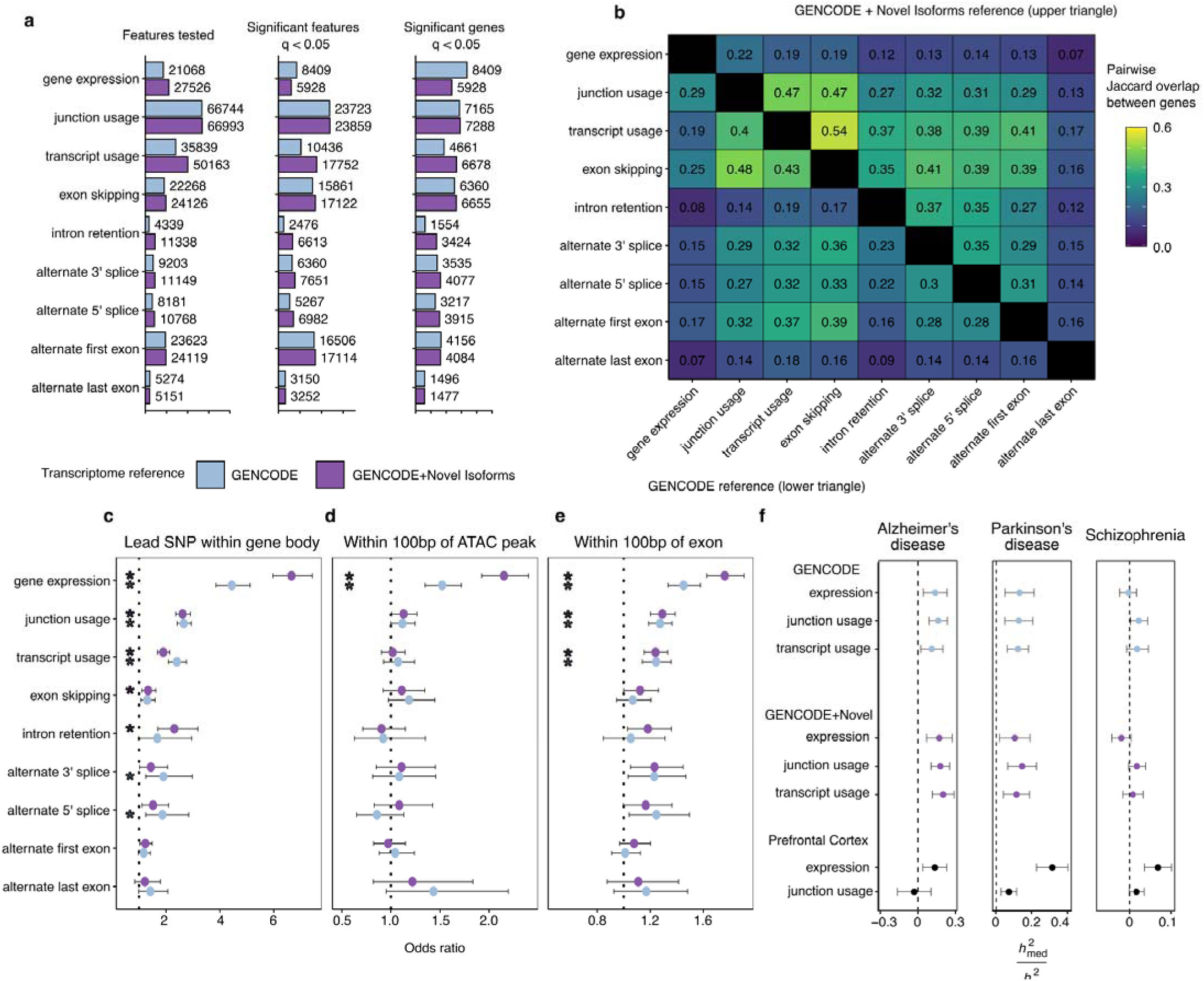
QTL mapping with augmented isoform reference. **a**, Discovery rate of each QTL type in either the GENCODE isoform reference (orange) of the augmented GENCODE + Novel Isoform reference (green). **b,** Pairwise sharing of genes with significant QTLs between each pair of QTL modalities in either the GENCODE reference (lower triangle) or the augmented GENCODE+Novel Isoform reference. **c-e**, Enrichment tests (two-sided Fisher’s exact test) comparing locations of significant lead QTL SNPs for each feature type (q <0.05) compared to null SNPs (q > 0.05). * refers to P < 0.05 after Bonferroni correction for multiple testing. **f**, Estimated proportion of heritability mediated by cis-genetic component of expression, junction usage or transcript usage (□^2^med/□^2^g) in three diseases by microglia and bulk dorsolateral prefrontal cortex.

We then compared the overlap between our different types of QTL modality across genes. By including the additional novel isoforms we find a greater degree of sharing between splicing QTL types, particularly between intron retention QTLs (**Figure 4b**). We compared the enrichment of our lead QTL SNPs in different genomic features, including gene bodies (the coordinates including the start and end of each gene) (**Figure 4c**), microglia-specific ATAC-seq peaks (**Figure 4d**), and exons (**Figure 4e**). We observed enrichment within the gene body of most QTL modalities, whereas only eQTLs were enriched in ATAC-seq peaks, and eQTLs, tuQTLs and juQTLs were enriched within exons.

### Novel isoforms explain new colocalizations with disease GWAS

We first applied our sets of expression and splicing QTLs to explain disease heritability using mediated expression score regression (MESC) (Yao et al. 2020) (**Figure 4f**). MESC estimates the proportion of common variant disease heritability (□^2^_g_) that is mediated through a set of QTLs (□^2^_med_ ). We found evidence of non-zero mediation of heritability by microglia expression and splicing QTLs in the two neurodegenerative diseases Alzheimer’s disease and Parkinson’s disease, but not the neuropsychiatric disease Schizophrenia, in line with previous studies in myeloid cells (Towfique Raj et al. 2014; Nott et al. 2019; Novikova et al. 2021). In contrast, expression and splicing QTLs generated in bulk brain cortical samples (Y. I. Li et al. 2019) mediated non-zero heritability of all three diseases, with the highest mediation of Parkinson’s disease heritability observed in bulk brain eQTLs.

We then colocalized our sets of microglia QTLs with individual genome-wide significant loci in Alzheimer’s disease (Bellenguez et al. 2022) (**Figure 5**) and Parkinson’s disease (Nalls et al. 2019) (**Figure 6**) to identify new mechanisms of risk mediation by effects on isoform usage and splicing (**Supplementary Table 10**). In Alzheimer’s disease we identified between 32 and 37 of the 78 examined risk loci to contain colocalized expression or splicing QTL types at a posterior probability > 0.5 (**Figure 5a**). We then compared the maximum colocalization probability in each locus with expression and splicing to determine whether the evidence favored an expression-based mechanism vs a splicing mechanism (**Figure 5b**). We observed the previously published CD33 and MS4A6A loci, and for the first time the TREM2 and ACE loci to have stronger probabilities of splicing rather than expression, whereas our previously examined BIN1 and ECHDC3 loci favored gene expression, in line with previous work (Nott et al. 2019; Lopes et al. 2022; Kosoy et al. 2022) showing that the likely causal variants lie within microglia-specific enhancers for *BIN1* and *USP6NL* respectively. However multiple loci, including PLCG2, CTSH, and WDR81 had equally strong probabilities of being mediated by expression as by splicing. We show the 14 AD loci with at least 1 sQTL colocalization with a posterior probability > 0.9 of being mediated through splicing (**Figure 5c**). We observed that only 3 of the loci had corresponding chromatin accessibility QTL colocalization, further supporting a splicing mechanism between them. In each locus, we summed the number of different sQTL modalities. For example, in the well-studied CD33 locus, 5 different types of sQTL colocalized using the GENCODE-only reference, whereas using the augmented GENCODE+Novel reference retained 3 of these associations, renaming a Leafcutter juQTL from *CD33* to overlapping both *CD33* and the *CD33-SIGLEC22P* readthrough fusion (**Supplementary Fig. 10**), as well as a second colocalization with *NR1H2.* The vast majority of CD33 sQTL colocalizations involve the well-known exon 2 splicing as well as the previously reported retention of the first intron (Malik et al. 2015), giving us confidence that our sQTL colocalizations can uncover new insights into the disease-linked genetic regulation of splicing.

**Fig. 5.**
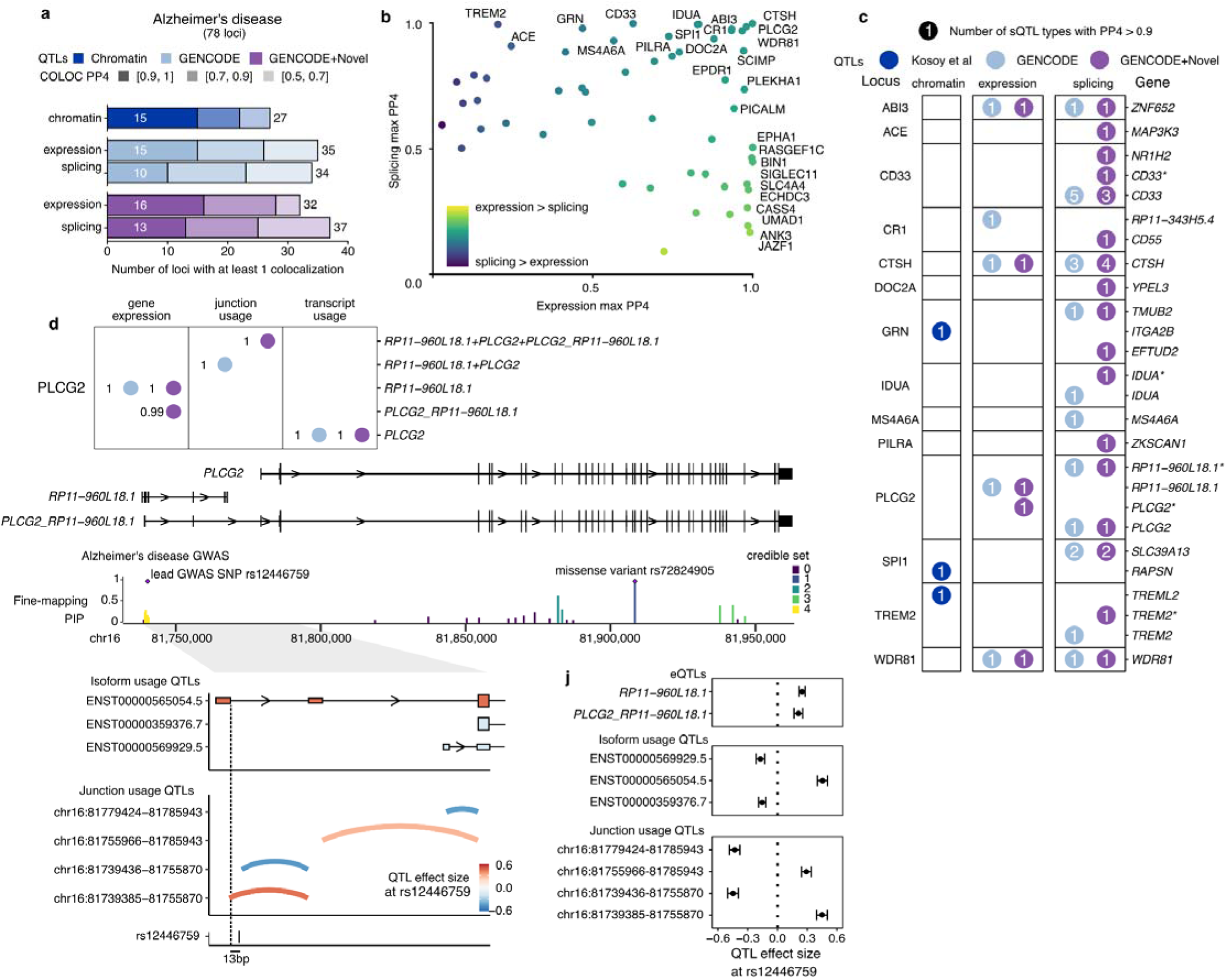
Novel isoforms improve interpretation of Alzheimer’s disease colocalization. **a**, Colocalization discovery rate for microglia chromatin, expression and splicing QTLs in Alzheimer’s disease at different thresholds of colocalization posterior probability (PP4). “Splicing” represents a combination of all 8 sQTL modalities. **b**, Comparing the maximum expression and splicing PP4 for each locus across both references. Labels refer to loci with PP4 > 0.9. **c**, Colocalization results at each locus in Alzheimer’s disease. Numbers within circles refer to the number of different sQTL modalities with a colocalization PP4 > 0.9. Asterisks refer to additional genes being implicated in fusion isoforms and/or overlapping splice junctions. **d**, Multiple QTLs colocalize with the PLCG2 Alzheimer’s disease risk locus. The lncRNA *RP11-960L18.1* upstream of *PLCG2* produces fusion isoforms detectable as novel isoforms in microglia. Fine-mapping of the *PLCG2* locus identifies a credible set of SNPs nearby the first exon of RP11-960L18.1, distinct from the rare missense mutation in the gene body of *PLCG2*. Transcript usage QTLs colocalizing with the GWAS variant show increased usage of the upstream TSS. **i**, Junction usage QTLs identify changes in splicing around the upstream fusion TSS and decreased usage of the downstream *PLCG2* TSS. **j,** Quantified effect sizes of each QTL type relative to the lead GWAS SNP rs12446759.

**Fig. 6.**
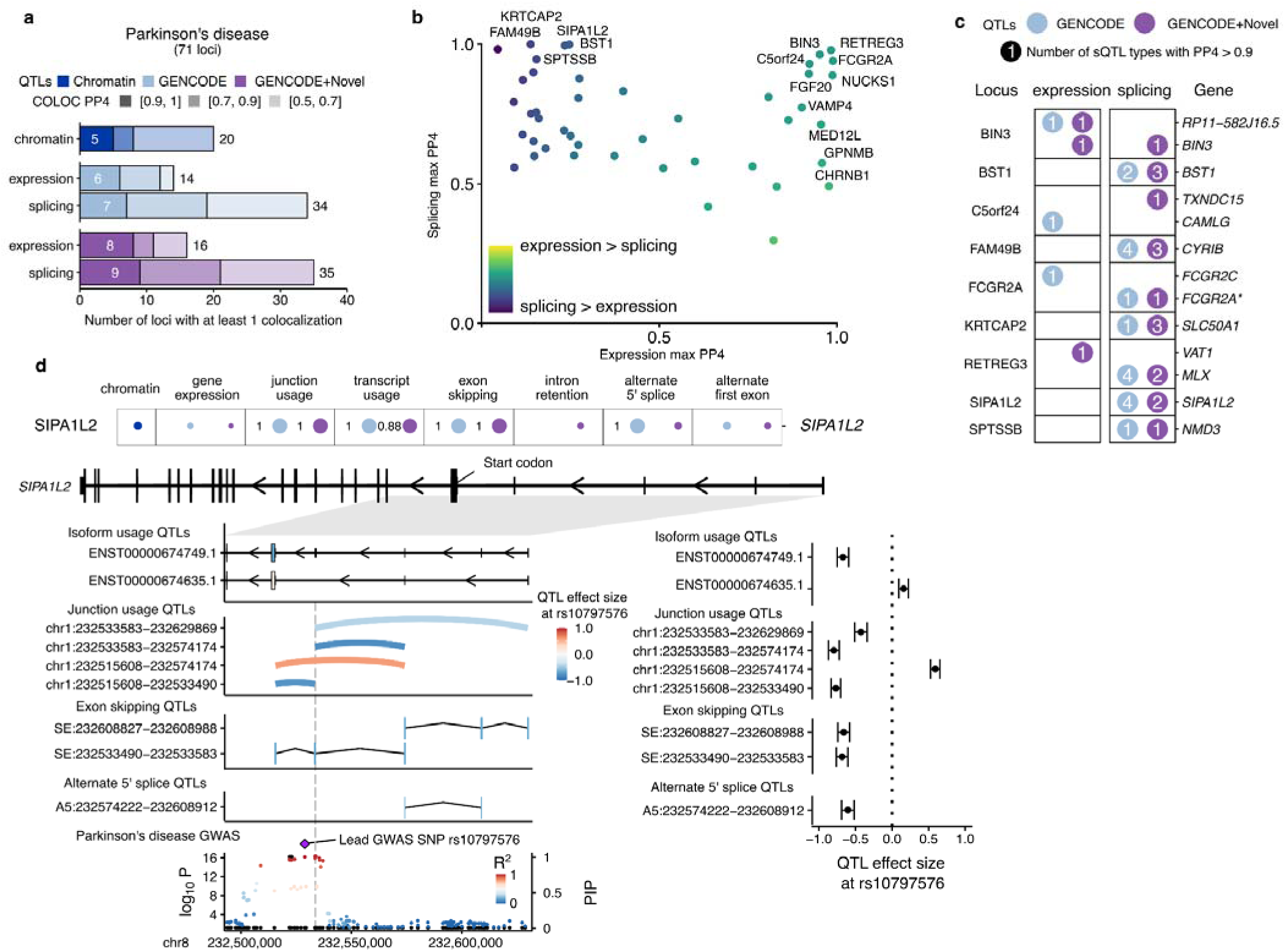
Splicing QTLs in Parkinson’s disease identifies 5’UTR exon in *SIPA1L2*. **a**, Colocalization discovery rate for microglia chromatin, expression and splicing QTLs in Alzheimer’s disease at different thresholds of colocalization posterior probability (PP4). “Splicing” represents a combination of all 8 sQTL modalities. **b,** Comparing the maximum expression and splicing PP4 for each locus across both references. Labels refer to loci with PP4 > 0.9. **c,** Colocalization results at each locus in Parkinson’s disease with at least one colocalized sQTL at PP4 > 0.9. Numbers within circles refer to the number of different sQTL modalities with a colocalization PP4 > 0.9. Asterisks refer to additional genes being implicated in fusion isoforms and/or overlapping splice junctions. **d,** Splicing QTL colocalizations at the SIPA1L2 locus implicate a non-coding cassette exon in the 5’UTR. Colocalization posterior probabilities (PP4) in each QTL modality in the two references. Values without numbers are PP4 <0.5. Isoform usage, junction usage and splicing event QTLs converge on the difference in inclusion of fourth 5’UTR exon upstream of the start codon (left panels) with effect sizes relative to the lead GWAS SNP in the SIPA1L2 locus (right panels). The SIPA1L2 GWAS locus in Parkinson’s disease contains multiple SNPs in high LD overlapping or nearby exon 4 (vertical dotted line). Black dots denote fine-mapping PIPs (right axis), whereas coloured dots refer to the -log_10_(P-value) of the GWAS (left axis). PIP: Posterior inclusion probability.

We further explored the *PLGC2* locus, where we observed colocalization with gene expression, junction usage and isoform usage (**Figure 5d**). *PLCG2*, a known interactor of *TREM2* (Andreone et al. 2020) was previously associated with AD risk through a rare missense coding variant (Sims et al. 2017), and was recently identified in the latest AD GWAS (Bellenguez et al. 2022) to have common risk variants, the lead SNP rs12446759 sitting upstream of the canonical *PLCG2* transcription start site, overlapping a long noncoding RNA *RP11-960L18.1* (**Figure 5e**). In our long-read RNA-seq data we observed multiple readthrough fusion isoforms which contained sequences from *RP11-960L18.1* splicing into *PLCG2*. A minor isoform in GENCODE, ENST00000565054.5 shares some of these exons but does not extend into the full coding sequence of PLCG2, whereas our long-read data identified novel isoforms containing the full exon content of both genes. We performed statistical fine-mapping of the *PLGC2* locus, identifying 5 independent credible sets of variants, the largest of which clustered around the lead common SNP rs12446759 (**Figure 5f**). We next examined the direction of effect of each QTL type relative to increasing dosage of the minor allele of rs12446759. *RP11-960L18.1* and the *PLCG2_RP11-960L18.1* fusion isoforms were increased in expression, whereas isoform usage QTLs showed an increased usage of the known fusion isoform ENST00000565054.5, and a relative decrease of two annotated isoforms using the canonical PLCG2 transcription start site. Finally, juQTLs found a complex pattern of intron usage, with the strongest effect size coming from a 5’ splice site change in the first exon of *RP11-960L18.1/PLCG2_ RP11-960L18.1.* Notably this splice site is 13bp from the lead GWAS SNP rs12446759, and 36bp from the lead junction usage QTL SNP rs12446781, immediately suggesting that the minor allele directly alters splice site choice. We used the deep learning models spliceAI (Jaganathan et al. 2019) and Pangolin (T. Zeng and Li 2022) which produce *in silico* predictions of variant effects on splice site usage. Although the scores for each variant are small (spliceAI: rs12446759-G-A donor gain = 0.03; rs12446781-G-C donor gain = 0.08; Pangolin: rs12446759-G-A splice gain = 0.04; rs12446781-G-C splice gain = 0.02), the two variants are in LD and may act together multiplicatively. Understanding how the splice site change is linked to changes in *RP11-960L18.1* and *PLCG2* expression warrants further biological experimentation.

In Parkinson’s disease, we observed more colocalizations (34-35) in sQTLs than eQTLs (14-16) in both isoform references (**Figure 6a**). Unlike in Alzheimer’s disease, we observed no PD risk loci with high eQTL colocalization that did not also have moderate evidence of also acting through splicing (**Figure 6b**), and several loci with high sQTL colocalization with little eQTL evidence, including the previously studied *FAM49B/CYRIB* locus (Navarro et al. 2021), *BST1*, and *SIPA1L2.* We looked further into the *SIPA1L2* gene, as it had 4 sQTL types colocalizing with at PD risk PP4 > 0.9 in the GENCODE reference, including junction usage, transcript usage, exon skipping and alternate 5’ splice site usage, and 2 sQTLs in the augmented reference (**Figure 6d**). Each sQTL type converged on a region of *SIPA1L2* upstream of the start codon in exon 5 (**Figure 6e**), and suggested that the minor allele of the lead GWAS SNP rs10797576 is associated with increased skipping of exon 4, which is part of the *SIPA1L2* 5’UTR. This has been previously identified as a junction usage QTL in IPS-derived sensory neurons (Schwartzentruber et al. 2018). The lead GWAS SNP and the lead sQTL SNPs are in high linkage disequilibrium (R^2^>0.99) all sit nearby the altered exon 4, with the closest SNP rs16857578, the lead junction usage QTL SNP being 5bp from the 5’ splice site. The T allele of rs16857578 is suggested by both spliceAI and Pangolin to cause a weakening of the 5’ splice site (SpliceAI donor loss delta score = 0.28; Pangolin splice site loss score = 0.78). As the skipping of exon 4 would not be predicted to alter the coding sequence of *SIPA1L2* and no eQTL was colocalized, the consequences of exon 4 splicing for the SIPA1L2 protein are unclear.

## Discussion

In this study, we present the isoMiGA, a large-scale RNA isoform atlas in human microglia built from 89 million long reads across 30 samples. We identified 35,879 novel isoforms and 2,238 novel genes, expanding our insight into the microglial transcriptome. In addition, we provide an expanded catalog of eQTLs and sQTLs that comprehensively characterizes the genetic regulation of gene expression and splicing in microglia.

In addition to increasing the number of potential protein-coding isoforms, our novel isoform catalog adds new categories of non-coding regulatory isoforms, such as intron retention, antisense, and readthrough fusions. For the well-studied AD risk genes *CD33* and *TREM2*, we observed multiple fusion isoforms to be highly expressed. Fusion isoforms have the potential to create new regulatory sequences, such as a new 5’UTR seen in *CD33*, or may be used to regulate translation through NMD. Using our expanded isoform reference, we observed that a fraction of the novel genes and isoforms respond to pro-inflammatory stimulation and show region specificity, suggesting that these new transcripts have specific functional roles. By quantifying full-length isoforms in single cells (Joglekar et al. 2021; Volden and Vollmers 2022; Al’Khafaji et al. 2023), future work could further our understanding of the relationship between isoform expression and the multiple states that microglia can occupy, particularly those most relevant to neurodegeneration, such as response to amyloid and tau pathology (Olah et al. 2020).

Our QTL catalog combines three published microglia cohorts with 121 additional samples generated for this study in a multi-ethnic meta-analysis, maximizing our discovery power in the largest such analysis to date. Furthermore, we have increased our ability to identify genetic effects on splicing by mapping multiple types of sQTL, rather than solely through junction usage. Although we observed that isoform usage, junction usage, and splicing event QTLs converge on the same genes, in a given locus they provide complementary results, in line with previous work (Garrido-Martín et al. 2021; Qi et al. 2022).

Combining our QTLs with the latest AD and PD GWAS have allowed us to identify, with high confidence, potential risk genes for 28 loci in AD and 15 in PD. We observed a clear demarcation between the 2 diseases in the relationship between genetic effects on expression and splicing in microglia. In AD, the predominant associations are seen in eQTLs (JAZF1, BIN1, ECHDC3, CASS4), or in loci with both eQTLs and sQTLs (PLCG2, CTSH). In comparison, PD risk loci either share eQTLs and sQTLs (BIN3, FCGR2A) or are exclusively mediated by sQTLs (SIPA1L2, FAM49B/CYRIB). As sQTL variants acting on splice sites will likely alter splicing in every cell-type in which that gene is expressed, sQTLs are by their nature less cell-type specific than eQTLs that act through a particular transcription factor-enhancer-promoter relationship. As the link between PD and microglia is less clear than in AD, it is likely that more PD loci will be identified in other CNS cell types.

We recognise that our study has several limitations. The correlation between short and long-reads at the isoform level is imperfect, and isoform assembly for long reads is still an ongoing challenge in the field. Furthermore, estimating isoform abundance in short reads using pseudoalignment is vulnerable to error with increasing levels of isoform length and complexity. We expect that increasing throughput and decreasing cost of long-read sequencing will allow future studies to characterize isoform abundance without the need for short reads. Secondly, we observed very little peptide support for our novel isoforms, likely due to trypsin digestion in shotgun proteomics disfavouring the creation of peptides that span exon junctions (Wang et al. 2018). Thirdly, our use of meta-analysis compared to having a single large cohort for QTL mapping puts a lower limit on the minor allele frequency of tested variants, which limits discovery to more common variants with potentially smaller effect sizes, and also makes assumptions about the sharing of effects across each cohort. Finally, colocalization between QTLs and GWAS imply evidence of shared genetic effects but does not directly test for causality. Unlike eQTL variants, which are now widely tested with massively parallel reporter assays (Cooper et al. 2022), genetic validation of splicing variants, particularly those that fall outside of exons or splice sites, remain challenging to validate experimentally or computationally.

In summary, we have generated both an isoform atlas and an updated catalog of genetic effects on the human microglial transcriptome, which we use to derive insights into specific loci associated with AD and PD. Our findings represent ongoing work in the field to better characterize an important causal cell-type in neurodegeneration, and will generate new mechanistic insights in the field.

## Methods

### Human brain tissue

Post-mortem brain samples were obtained from the Netherlands Brain Bank (NBB), the Neuropathology Brain Bank and Research CoRE at Mount Sinai Hospital, NY, Rush University Medical Center/Rush Alzheimer’s Disease Center in Chicago, IL and the Mount Sinai/JJ Peters VA Medical Center NIH Brain and Tissue Repository in the Bronx, NY. Rush Alzheimer’s Disease Center samples were collected as part of two prospective studies of aging: the Religious Orders Study and the Memory and Aging Project (ROSMAP). All persons in ROSMAP agree to annual clinical evaluation and organ donation at death. Both studies were approved by an Institutional Review Board of Rush University Medical Center. Participants signed informed and repository consents and an Anatomic Gift Act. The permission to collect human brain material was obtained from the Ethical Committee of the VU University Medical Center, Amsterdam, The Netherlands, and the Mount Sinai and Mount Sinai/JJ Peters VA Medical Center Institutional Review Boards (IRB). Informed consent for autopsy, the use of brain tissue and accompanied clinical information for research purposes was obtained per donor ante-mortem. All autopsies were performed with written consent from the legal next-of-kin. The study was performed under IRB-approved guidance and regulations to keep all patient information strictly de-identified. All research conformed to the principles of the Helsinki Declaration. Detailed information per donor, including tissue type, age, sex, postmortem interval, and pH of cerebrospinal fluid is provided for the long-read samples (**Supplementary Table 1)**, and newly generated short-read samples (**Supplementary Table 3**). Participants did not receive compensation.

### Microglia isolation

This study combined samples from two different protocols for microglia isolation. The Raj lab employed magnetic sorting with CD11b beads (Lopes et al. 2022) on four distinct brain regions: the medial frontal gyrus, superior temporal gyrus, thalamus, and subventricular zone. The Roussos lab employed fluorescence-activated cell sorting with antibodies against CD45 and CD11b (Kosoy et al. 2022) exclusively on samples of the prefrontal cortex. For long-read sequencing, the Raj lab isolated 10 samples from 7 donors from the medial frontal gyrus and subventricular zone, and the Roussos lab isolated 20 samples from 20 donors (**Supplementary Table 1**). For short-read sequencing, to augment the published data, the Raj lab generated an additional 37 samples, and the Roussos lab an additional 84 (**Supplementary Table 2**).

### Long-read RNA sequencing

Each sample was sequenced on a Pacific Biosciences Sequel II using a single-molecule real-time (SMRT) flow cell 8M for each sample individually. Raw movie files were collapsed into circular consensus sequences using PacBio CCS (v4.0). Barcodes were then removed with LIMA (v1.11) and trailing polyA and polyT tails were clipped using PacBio isoseq3 refine (v3.2), to create sets of full-length non-concatemer (FLNC) sequences, yielding on average 29.7M reads per sample (range 6.4-39.5). FLNC reads were then aligned to the hg38 reference genome using pbmm2 (v1.4.0), the PacBio implementation of minimap2 (H. Li 2018). Unique alignment rates were > 99.5% for all samples. Each aligned sample was assembled into isoforms using Stringtie2 (v2.2.1) (Kovaka et al. 2019; Shumate et al. 2022) in hybrid assembly mode, combining both long- and short-read RNA-seq reads for each sample using the --mix argument and excluding mono-exonic (unspliced) isoforms. Each sample’s isoform assembly was then merged using stringtie --merge and expression of each transcript in each sample was then re-quantified using stringtie --estimate. Isoforms were compared to the GENCODE v38 reference annotation to determine novelty. All annotated isoforms were then kept. Any novel transcript had to be identified in at least 2 samples, resulting in 93,199 annotated and 42,153 novel isoforms. Isoforms were then further processed using SQANTI (v3) (Tardaguila et al. 2018), a software toolkit which further refines and classifies isoforms using the same GENCODE v38 reference annotation (GENCODE v38). All isoforms assigned as a full splice match, as in exactly matching a GENCODE isoform, were kept. All novel isoforms had to pass the following criteria:

- No splice junctions must be predicted to be from reverse transcriptase (RT)-switching.
- Isoforms with an annotated transcript termination site (TTS) were kept.
- If a novel TTS is used, due to the likelihood of novel TTS arising from intrapriming of adenosine-rich genomic DNA or cDNA, the immediate 20bp of genomic sequence downstream of the TTS must meet all three criteria:

- Fewer than 6 adenosines in a row directly downstream
- An overall adenosine content less than 60%
- Roy & Chanfreau (Roy and Chanfreau 2020) score < 15. This counts adenosines in the downstream 19bp, counting any adenosines in the first 6bp twice.

This resulted in a final set of 128,436 isoforms, of which 35,879 were novel. The filtering most affected isoforms classified as genic, antisense, and intergenic, whereas novel in catalog isoforms were the least affected (**Supplementary Fig. 3**). The novel isoforms were then appended to the GENCODE v38 set of annotated isoforms to form a combined set (GENCODE+Novel). Open reading prediction was performed with GeneMarks (v5.1) which is included with SQANTI.

As a comparison, we ran Bambu (v3.0.5) (Chen et al. 2023), an alternative transcript discovery tool for long-read sequencing data, using the default settings and identical downstream filtering. Bambu discovered 124,071 isoforms, of which 19,145 were novel. We compared the Stringtie2 and Bambu transcript sets using Gffcompare (Pertea and Pertea 2020) and found 4,725 novel isoforms and 70,365 annotated isoforms exactly matched between the two references. Although the exact replication rate for novel isoforms was low, we note that the two underlying assembly tools use different models and assumptions.

### Short-read RNA sequencing

All short read samples were uniformly processed using RAPiD, a Nextflow pipeline for uniform processing and quality control of RNA-seq data. All samples were first aligned to the hg38 genome build with STAR (Dobin et al. 2013) (v.2.7.2a). Expression of either the GENCODE or GENCODE+Novel isoform sets were estimated in both short and long-read RNA-seq samples using Salmon (Patro et al. 2017) (v1.4). Indexes were constructed using the FASTA sequences of the transcript sets. Quantification was then run with the following options: selective alignment, sequence specific bias model, fragment-level GC bias model, and positional bias model. Transcript abundances were summarized as both estimated read counts and normalized transcripts per million (TPM). Total gene expression was calculated by tximport (v1.24) (Love, Soneson, and Robinson 2017), which sums transcript expression in counts or TPM from each transcript for each gene.

### Induced microglia cells

Isogenic induced pluripotent stem cell (iPSC) lines for a single donor with *LRRK2* G2019S wild-type, heterozygous, and homozygous variants were provided by the Cookson Lab (NIH). Genotype was validated by Sanger sequencing, and all clones displayed a normal karyotype and standard pluripotent expression markers. iPSC lines were differentiated to induced hematopoietic stem cells (HPCs) over a period of 11-15 days, and subsequently to microglial cells (iMGLs) for 25 days following a published protocol (Abud et al. 2017). Each HPC to iMGL differentiation is considered one biological replicate. Mature iMGLs were plated at 500,000 cells per well of a 6-well plate and stimulated for 24 hours with LPS-EK ultrapure (InvivoGen) 10 ng/ml, IFNg (R&D Systems) 20 ng/ml or baseline (media + PBS). After stimulation cells were washed with PBS and pellets were resuspended in 350 µl of RLT + 1% 2-Mercaptoethanol (Sigma Aldrich) and stored at -80°C. RNA was isolated using the RNeasy Mini kit (Qiagen) following the manufacturer’s instructions including optional DNase treatment. RNA was stored at -80 °C prior to library preparation. Library preparation and sequencing was performed at Genewiz Inc. using the SMART-Seq v4 Ultra Low input library preparation protocol, which uses poly-A selection. Samples were sequenced with a depth of 30 million 150-bp paired-end reads using Illumina HiSeq 4000 platform.

### Differential analyses of region and stimulation

Both the stimulated and region-specific microglia cohorts were processed using the same quality control and alignment pipelines. For each cohort, isoform-level expression of either the GENCODE v38 or GENCODE+Novel transcriptome references was quantified with Salmon. Differential expression of iMGLs in response to stimulation with LPS and IFNg was performed using DESeq2 (v1.30.1) and edgeR (v3.32.1) on the estimated count matrices, filtering lowly expressed genes with a median TPM < 0.1 and including % mRNA bases, as estimated by Picard, as a covariate in the model. As the 3 genotype groups were balanced across the stimulation conditions, we did not include it as a covariate. For the region-specific analysis of the Raj postmortem microglia, DREAM (v1.2) (Hoffman and Roussos 2021) was applied to account for repeated donors, adjusting for sex, age, batch, donor id, cause of death, % mRNA bases, median insert size, and % ribosomal bases. Differential transcript usage (DTU) analysis was performed using satuRn (v1.2) (Gilis et al. 2021). Filtering was performed using edgeR (v3.36) (Robinson, McCarthy, and Smyth 2010) where the minimum count required for the minimum group size was 150 and the minimum proportion of samples in the smallest group that expresses the gene is 0.70. For both cohorts, the same covariates were applied for DTU analysis, with the exception of donor ID in the region analysis, as SatuRn cannot account for repeated donors.

### Quantitative trait locus mapping

#### Single cohort QTL mapping

We prepared six separate cohorts of paired genotype-phenotype data from human microglia, including the Gaffney, Roussos and Raj data, splitting the Raj samples by brain region of origin into four cohorts. We first mapped expression QTLs in each cohort individually using tensorQTL (Taylor-Weiner et al. 2019) (v.1.0.2) as previously described (Lopes et al. 2022), optimizing correction for technical variation by varying the number of estimated PEER factor covariates (Stegle et al. 2012) between 0 and 30 in steps of 5 and picking the number of covariates that maximized eQTL discovery. This suggested an optimum PEER factor number of 5 for each Raj cohort and 10 for Roussos and Gaffney.

#### Multi-cohort QTL meta-analysis

To combine all six cohorts in a random-effects meta-analysis, we constructed a new pipeline based on mmQTL (v26a), a linear mixed modeling framework which allows for both multiple ancestries and repeated donors (B. Zeng et al. 2022). Briefly, for each of the six cohorts, phenotype data was first prepared and normalized (see below), PEER factors estimated and regressed out. Genotype data was converted into Plink (v2.3) format and a genotype relatedness matrix constructed using GCTA (Yang et al. 2011) (v1.93.2). Lists of phenotypes were harmonized between cohorts by allowing any phenotype that is present in at least 2 of the 6 cohorts to be put forward for QTL mapping. QTL mapping was then performed for all SNPs within 1 megabase either side of each phenotype’s coordinates. mmQTL then performs a random-effects meta-analysis, accounting for shared donors by comparing SNPs with null association. The random effects P-values for each phenotype-SNP pair are then corrected twice, for the number of SNPs using Benjamini-Hochberg adjusted and then the number of features using Storey’s q value.

#### Phenotype preparation

Lowly expressed genes or isoforms are commonly filtered out of QTL mapping studies using a minimum expression filter. Because TPM values by definition sum to 1,000,000, and the combined GENCODE+Novel reference includes additional genes and isoforms, we adjusted the minimum TPM threshold to capture similar numbers of annotated genes, using a median TPM > 1 in the GENCODE reference and a median TPM > 0.1 the combined reference. Gene expression: Counts and TPM values for each gene were used. For each TPM-filtered gene, the corresponding counts were then TMM normalized using limma voom (Law et al. 2014) (v3.46.0). Transcript usage: After TPM filtering, transcript TPMs in each sample were divided by the sum of all isoforms for that gene. Splice Junction usage: All splice junctions from all samples were then clustered together using leafcutter_cluster_regtools.py from Leafcutter (Y. I. Li et al. 2018) (psi_2019 branch) on default settings. Junction counts were converted to counts per million (CPM) by dividing by the total junction counts per sample and multiplied by 1 million. Any junction with median CPM < 1 was removed. Splicing event usage: For each transcript reference, splicing events were constructed using SUPPA (Trincado et al. 2018) (v2.3) prepareEvents, pooling isoforms with overlapping exons. Then for each transcript TPM matrix, percent-spliced-in (PSI) values for each event were quantified using SUPPA quantifyPSI.

#### Normalization of features

For each phenotype, any feature with > 25% missing data was removed. Any remaining missing values were imputed with mean for that phenotype. Any phenotype with a standard deviation of 0 was removed. Individual expression or ratio values were then scaled and centered between samples, and quantile-normalized between individuals (Degner et al. 2012) before PEER factors were calculated and regressed out.

### Colocalization analysis

We used Coloc (Giambartolomei et al. 2014) (version 3.2-1) to test whether SNPs from different loci in the latest Alzheimer’s disease and Parkinson’s disease GWAS co-localized with our microglia QTLs. For each genome-wide significant locus, we extracted the nominal summary statistics of association for all SNPs within 1 Mb either upstream or downstream of the top lead SNP (2-Mb-wide region total). In each QTL dataset, we then extracted all nominal associations for all SNP–gene pairs within that range and tested for co-localization between the GWAS locus and each gene. To avoid spurious co-localization caused by long-range LD, we restricted our co-localizations to GWAS SNP–QTL SNP pairs where the distance between their respective top SNPs was ≤500 kb or the two lead SNPs were in moderate LD (r2 > 0.1), taken from the 1000 Genomes (Phase 3) European populations using the LDLinkR package (version 1.1.2) (Myers, Chanock, and Machiela 2020). For all splicing QTL types we followed the same approach but collapsed features (transcript, junction, event) to return only the highest posterior probability of colocalization for each gene in each locus.

### Plotting

All plots were created using ggplot2 (version 3.3.3) in R (v4.0.4), with ggrepel (v0.9.1), ggfortify (v0.4.11), patchwork (v1.1.1), ggtranscript (v0.99.9) and ggbio (version 1.38.0) for additional layers of visualization.

## Data availability

Long-read data: https://www.synapse.org/#!Synapse:syn52052829

Gaffney short-read cohort: https://ega-archive.org/datasets/EGAD00001005736

Raj short-read microglia samples: https://dss.niagads.org/datasets/ng00105/

Roussos short-read batch 1: https://doi.org/10.7303/syn26207321

Roussos short-read batch 2: https://www.synapse.org/#!Synapse:syn52052829

IMGL short-read: Gene expression omnibus GSE240907

Novel isoform GTF and FASTA: https://zenodo.org/record/8290956

Count matrices for all cohorts: https://zenodo.org/record/8291211

All QTL summary statistics: https://zenodo.org/record/8250771

## Code availability

Short-read RNA-seq pipeline: https://github.com/CommonMindConsortium/RAPiD-nf/

Genotype quality control pipeline: https://github.com/RajLabMSSM/Genotype_QC_Pipeline_2.0

Long-read RNA-seq pipeline: https://github.com/RajLabMSSM/isoseq-pipeline/tree/master

QTL preparation and meta-analysis pipeline: https://github.com/RajLabMSSM/mmQTL-pipeline

All code used to produce analysis for figures: https://github.com/RajLabMSSM/isoMiGA

## Acknowledgments

We thank the patients and families who donated material for these studies. We thank Elizabeth Tseng of Pacific Biosciences who provided important critical feedback on long-read analysis. This study was supported by the following National Institutes of Health grants: T.R., J.H, E.B., D.M., E.C. and B.Z.M: NIA U01-AG058635, NIA R21-AG063130, NIA R01-AG054005, NIA U01-AG068880, NIA RF1-AG065926, NIA R56-AG055824, NIA P30-AG066514, NINDS U54-NS123743, and NINDS R01-NS116006. P.R., R.K., B.Z., S.P.K, S.A., Z.S., G.E.H. and J.F.F.: NIA U01-AG058635, NIA R01-AG067025, NIA R01-AG082185, NIA R01-AG050986, NIA R01-AG065582, NIMH R01-MH125246, and NINDS U01-NS125580. A.M.G., A.M., and A.G.E.: NIA U01-AG058635.

This work was supported in part through the computational resources and staff expertise provided by Scientific Computing at the Icahn School of Medicine at Mount Sinai and supported by the Clinical and Translational Science Awards (CTSA) grant UL1TR004419 from the National Center for Advancing Translational Sciences. Research reported in this paper was supported by the Office of Research Infrastructure of the National Institutes of Health under award number S10OD026880 and S10OD030463. The content is solely the responsibility of the authors and does not necessarily represent the official views of the National Institutes of Health.

## Author Contributions

Study was conceived by JH, PR, and TR. Data analysis was led by JH, with contributions from EB, RK, BZ, AR, BZM, and C-FT. Data generation was performed by EC, DM, AGE, EN, GJLJS, AM, SPK, and JFF. The manuscript was written by JH, with input from all co-authors.

## List of Supplementary Materials

Supplementary Materials and Methods

Supplementary Text

Figs. S1 to S11

Tables S1 to S10

