## Supplementary Materials for "Long-read RNA-seq atlas of novel microglia isoforms elucidates disease-associated genetic regulation of splicing"

**Supplementary Material**

**Table of Contents**


**Supplementary Methods** **2**
External Datasets 2
RT-PCR of readthrough fusion isoforms 2
Mass Spectrometry 3
Fine-mapping analysis 4
Mediated expression score regression (MESC) 4
Genotyping quality control 5
**Supplementary Figures** **6**
Supplementary Figure 1 - Long-read RNA-seq processing pipeline. 6
Supplementary Figure 2 - Read length distribution in the IsoSeq samples 7
Supplementary Figure 3 - Quality Control of long-read RNA-seq 8
Supplementary Figure 4 - Correlating gene and isoform expression between cohorts 9
Supplementary Figure 5 - Mass spectrometry on human microglia 10
Supplementary Figure 6 - CD33 isoforms 11
Supplementary Figure 7 - RT-PCR confirmation of fusion isoforms in TREM2 and CD33. 12
Supplementary Figure 8 - Differential gene expression in IPS-derived microglia (iMGLs) in response to LPS treatment. 13
Supplementary Figure 9 - Predicted ancestries in each cohort. 14
Supplementary Figure 10 - eQTL discovery between the two references. 15
Supplementary Figure 11 - splicing QTLs in CD33. 16

### Supplementary Methods

#### External Datasets

##### *Gaffney Microglia Cohort*

We downloaded raw data of bulk RNA-seq for primary microglia samples from European phenome-Genome Archive (EGA) (Accession ID: EGAD00001005736). For each sample we downloaded the FASTQ files.

##### *Genome-wide association study (GWAS) data*

We downloaded full summary statistics and lists of genome-wide significant loci for the following diseases: Alzheimer's disease (AD) (Lambert et al. 2013; Bellenguez et al. 2022), Parkinson's disease (PD) (Nalls et al. 2019), and Schizophrenia (Trubetskoy et al. 2022). Missing fields in the nominal statistics were dealt with as follows: standard error was calculated from the effect size and P-value; minor allele frequency was taken from the European samples from 1000 Genomes; SNP coordinates or RS ID were matched using Ensembl (release 99). For Bellenguez AD GWAS we used the full summary statistics from the Stage 1 analysis, as the combined Stage 1&2 meta-analysis was not available. For PD we removed any loci that did not pass the final quality control filtering according to the column "Failed final filtering and QC". To avoid double-counting in colocalization, if multiple GWAS loci overlapped (within 1 megabase), we retained the locus with the lowest P-value. Due to the complex recombination structure within both regions, loci overlapping the human MHC/HLA region (hg19 chr6:28,477,797–33,448,354) or the MAPT H1/H2 haplotype region (hg19 chr17:43,628,944–44,571,603) were removed. When conditionally independent loci were listed, only the primary association was kept due to lack of conditional summary statistics.

##### *In silico splicing prediction*

Delta scores for spliceAI and Pangolin were pulled using the spliceAI lookup tool (https://spliceailookup.broadinstitute.org/).

#### RT-PCR of readthrough fusion isoforms

Human pluripotent stem cell-induced microglia and microglia freshly isolated from postmortem tissue were collected in cold PBS and centrifuged at 300xg for 5 minutes at 4°C. RNA was extracted with the Monarch Total RNA Miniprep Kit (New England BioLabs #T2010S), and cDNA synthesis was performed on 1 µg RNA using the High-Capacity cDNA Reverse Transcription Kit (Applied Biosystems #4368814) following the manufacturer's recommendations. Products were amplified using Q5® High-Fidelity DNA Polymerase (New England BioLabs #M0491L) in a thermocycler, using primers spanning the exon-exon junction for the novel fusion isoforms: CD33 Forward:

CTGCCATGGGCTAATCTCTC, CD33 Reverse: TCCTTCCACTCTGAGGTGCT; TREM2 Forward: TGAACACTGCTTCTCCTGGA, TREM2 Reverse GCCTGTGAGAATGGAGGGAT. Products were run on 2% agarose gels, purified with the NucleiSpin Gel and PCR Clean-up kit (Macherey-Nagel #740609) and Sanger sequenced to validate novel isoforms with the same primers used for amplification.

#### Mass Spectrometry

##### *Sample preparation*

The cell pellet was solubilized in 50 µL of lysis buffer, containing 5% SDS, 50 mM TEAB, pH 8.5 with the 1 mM PMSF, 2 µg/mL Aprotinin, 10 µg/mL Leupeptin, and 10 mM NaF, via sonication in the sonication bath with ice for 5 min, followed by gentle mixing of the lysates in a Thermomixer at ~300 rpm for 5 min at room temperature. The lysate was centrifuged at 15,000 x g for 10 min at room temperature, and the supernatant was transferred into a new tube, with its protein concentration measured using BCA Protein Assay (ThermoFisher Scientific). Proteins were reduced using 5 mM DTT for 45 min and alkylated using 10 mM iodoacetamide for 45 min in the dark; both incubation steps were carried out at 25 °C with gentle shaking (300 rpm) in a Thermomixer. The alkylated proteins were then digested using the S-Trap™ micro spin columns (PROTIFI) according to the manufacturer's instructions. Briefly, 1:1 Trypsin/LysC mix was used at 1:25 (w/w) enzyme/protein ratio and incubated at 25 °C overnight (~ 16 hr). After digestion, peptides were eluted, pooled, and dried in a SpeedVac. Dry peptides were resuspended in 0.1% formic acid (FA) and cleaned via SPE C18 on stage tip packed with 2 discs of Empore C18 material (Empore Octadecyl C18, 47 mm; Supleco, 66883-U); 50% acetonitrile (ACN), 0.1% FA was used for elution of peptides from the C18 stage tips after two washes with 100 µL of 0.1% trifluoroacetic acid (TFA). The peptide sample was adjusted to 0.1 µg/µL with 3% ACN, 0.1% FA containing 0.01% n-Dodecyl-beta-Maltoside for LC-MS/MS analysis.

##### *Data acquisition*

Five microliter of the peptide sample was injected for analysis by LC-MS/MS using an Orbitrap Fusion Lumos Tribrid mass spectrometer (Thermo Scientific) connected to a nanoACQUITY UPLC system (Waters) (buffer A: 0.1% FA with 3% ACN and buffer B: 0.1% FA in 90% ACN. Peptides were separated by a gradient mixture with an analytical column (75 µm i.d. × 20 cm) packed using 1.9-µm ReproSil C18 and with a column heater set at 48 °C. Peptides were separated by an LC gradient: 2-6% buffer B in 1 min, 6-30% buffer B in 84 min, 30-60% buffer B in 9 min, 60-90% buffer B in 1 min, and finally 90% buffer B for 5 min at 200 nL/min. The data-independent acquisition (DIA) MS/MS scan was performed in the by higher-energy collision dissociation (HCD) mode with the following parameters: precursor ions from 350-1,650 m/z were scanned at 120,000 resolutions with a maximum ion injection time of 50 ms and an automatic gain control (AGC) target of 4E5. The scan rage of m/z (isolation window) of DIA windows from

377 (54), 419(32), 448(28), 473.5(25), 497.5(25), 520.5 (23), 542.5 (23), 564.5 (23), 587 (24), 610.5 (25), 635 (26), 660 (26), 685.5 (27), 712.5 (29), 741 (30), 771 (32), 803.5 (35), 838.5 (37), 877 (42), 921 (48), 972 (56), 1034.5 (71), 1133.5 (129) and 1423.5 (453) were scanned at 30,000 resolution with an maximum ion injection time of 54 ms and an AGC target of 3E6. The isolated ions were fragmented by HCD with an energy level of at 30%.

##### *Proteomics data analysis*

The raw DIA data were processed by DIA-NN 1.8.1 (Data-Independent Acquisition by Neural Networks) (Demichev et al. 2020) and searched against a FASTA constructed from combining *Homo Sapiens* GENCODE v38 amino acid sequence and novel isoforms of long-read RNA expressed in microglia with a total of 149,893 proteins and common contaminants. The fasta file was *in silico* digested to generate a spectrum library. A peptide search was performed with trypsin as digestion enzyme allowing a maximum of two missed cleavages. Cysteine carbamidomethylation was set as a fixed modification; acetylation of protein N-terminal and methionine oxidation were set as variable modifications. Precursor false discovery rate (FDR) is set to 1% and used for further novel isoform filtering.

#### Fine-mapping analysis

We fine-mapped each locus of the latest AD GWAS (Bellenguez et al. 2022) with SuSiE (v0.11.92) (Zou et al. 2022). For each locus, we gathered all SNPs within 2-Mb windows (± 1 Mb flanking the lead GWAS SNP) and filtered out SNPs with a minor allele frequency (MAF) < 0.001. LD correlation matrices (in units of r) were obtained for each locus from the GWAS data, calculated by Bellenguez et al. Statistical fine-mapping was performed on each locus separately using SuSiE, a Bayesian method that 1) provides a posterior inclusion probability (PIP) that each SNP is causal from 0 to 1 and 2) provides credible sets (CS) of SNPs that have been identified as having a high PIP of being causal, which we have set at a threshold of PIP ≥ 0.95. We set the number of causal SNPs to five.

#### Mediated expression score regression (MESC)

We applied MESC (Yao et al. 2020) (v1.0) to genome-wide QTL summary statistics for gene expression, transcript usage and junction usage in the two microglia transcriptome references. As a comparison we applied MESC to expression and junction-usage QTLs generated from bulk brain RNA-seq (Li et al. 2019). For each SNP-feature association we created input files containing the coordinates of each feature (gene, transcript, junction), the total number of donors tested for that feature based on whether each of the six cohorts contributed to the meta-analysis, and the random effects Z-score estimated by mmQTL. We generated expression scores for each QTL set first, and then estimated the SNP heritability ($\hbar^2_g$) and mediated cis-genetic component ($\hbar^2_{med}$) using GWAS summary statistics for Alzheimer's (Bellenguez et al.

2022), Parkinson's (Nalls et al. 2019), and Schizophrenia (Trubetskoy et al. 2022), processed using munge_sumstats.py from LDSC (Bulik-Sullivan et al. 2015).

#### Genotyping quality control

Genotype data in VCF format was downloaded for the three cohorts. Any previously imputed variants were stripped out before imputation was performed using the HRC TOPMed imputation panel. Variants were removed for failing the following quality control criteria: minor allele frequency > 1%, Hardy-Weinberg equilibrium P > 1e-6. Ancestry and relatedness was estimated using Somalier (Pedersen et al., n.d.) (v0.2.12). Sample mismatches between RNA and DNA were checked using MBV from the QTLtools package (Fort et al. 2017) (v.1.3). Genotype samples with missing RNA-seq were removed. Any sample swaps identified by the matching of an RNA-seq sample with >80% heterozygous variant match to the "wrong" genotype were assumed to be the true pairing and incorporated into the sample metadata.

### Supplementary Figures

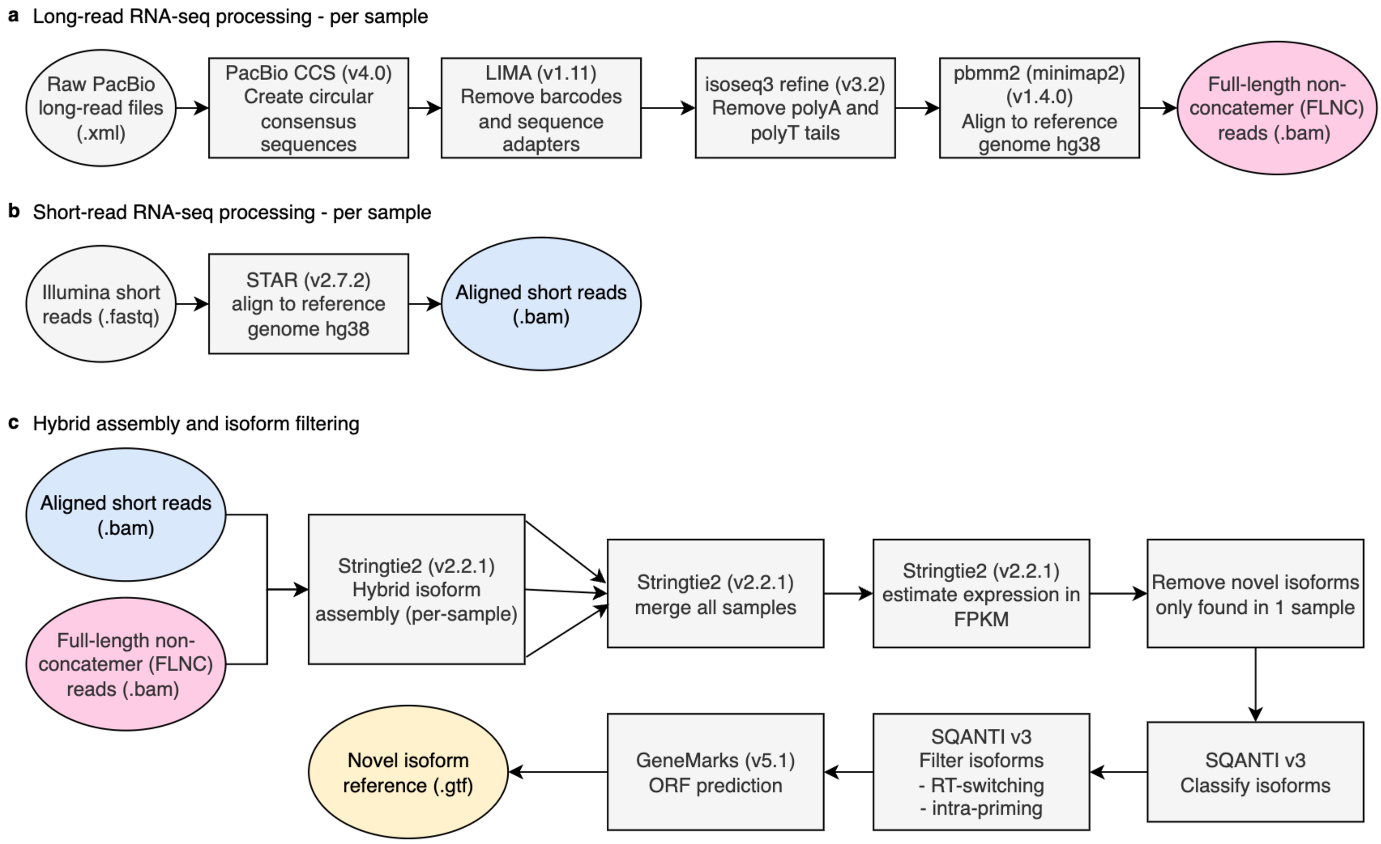


**Supplementary Figure 1 - Long-read RNA-seq processing pipeline.**

Flowchart created with www.draw.io

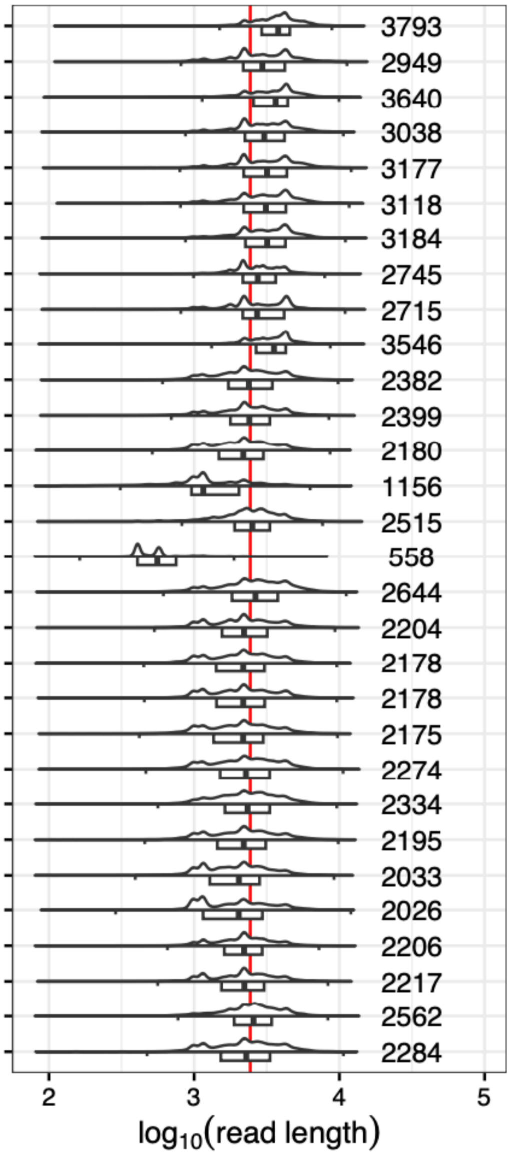


#### Supplementary Figure 2 - Read length distribution in the IsoSeq samples

a) Distribution of lengths in bases or 10,000 randomly sampled mapped reads in each sample. Length in base pairs. Boxplots show the first quartile, median and third quartile of the distribution. Whiskers extend to 1.5 times the interquartile range. Violin plots show the density of the distribution with each sample at a fixed width. Horizontal lines show the range of the distribution. Labels report median read length in each sample. Red vertical line at 2433bp denotes the median of all sampled reads.

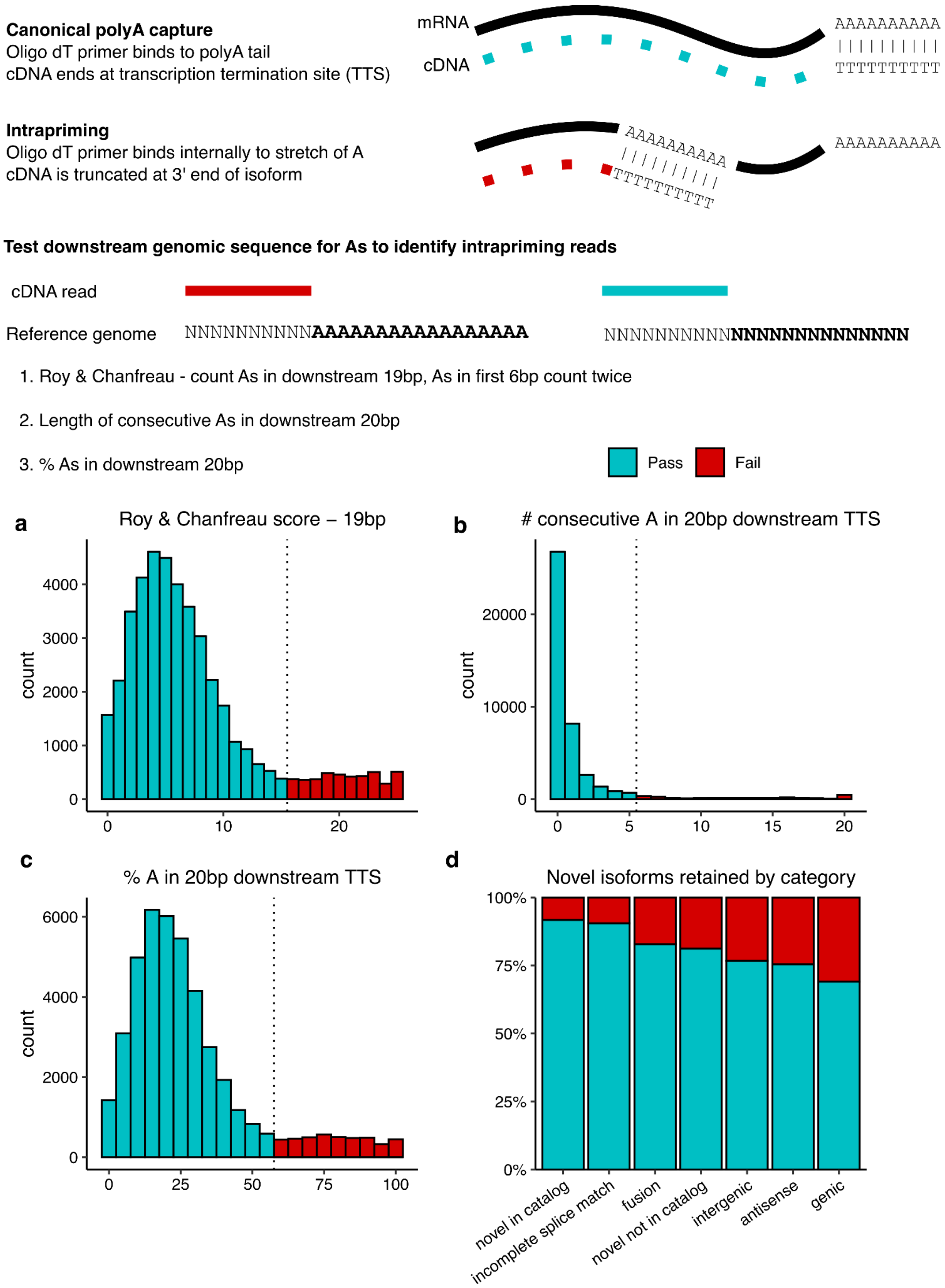


#### Supplementary Figure 3 - Quality Control of long-read RNA-seq

All novel isoforms are subjected to QC to remove spurious truncated isoforms resulting from intrapriming - where the oligo dT primer binds to an internal stretch of Adenosine-rich RNA. The likelihood of intrapriming can be estimated by examining the genomic DNA sequence downstream of the transcription termination site (TTS). **a-c**) Distributions of the 3 scores for A content in all novel microglia isoforms. Dotted line refers to the cutoff for removal of likely intrapriming artifacts. **d)** The proportion of each novel isoform type removed by filtering.

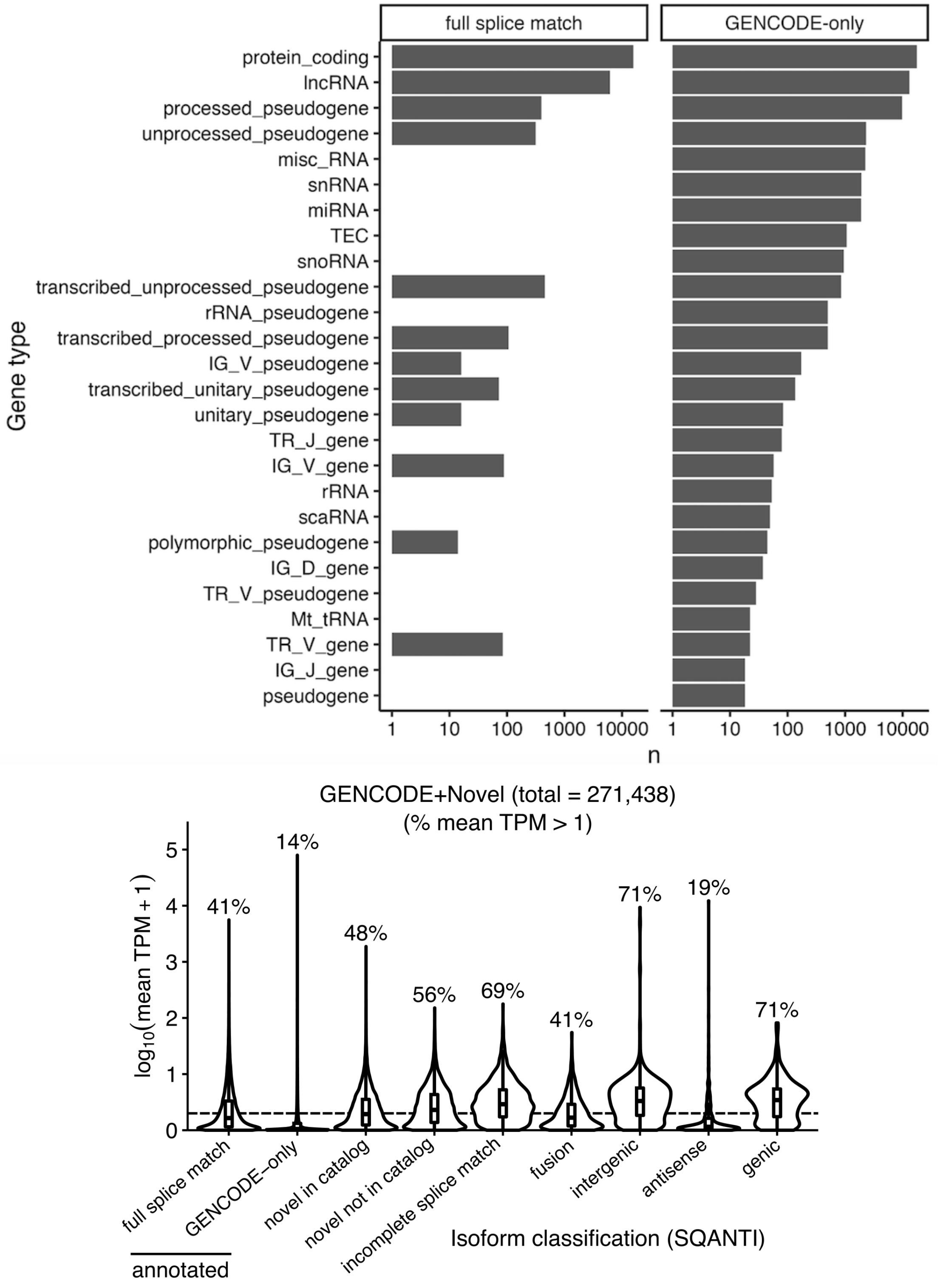


**Supplementary Figure 4 - Correlating gene and isoform expression between cohorts**

Upper panel: Counts of gene types for annotated genes found in long-read vs GENCODE-only genes. Only displaying gene types with > 5 genes in each category.

Lower panel: Distribution of short-read RNA-seq expression values for each isoform type. Dotted line denotes TPM=1.

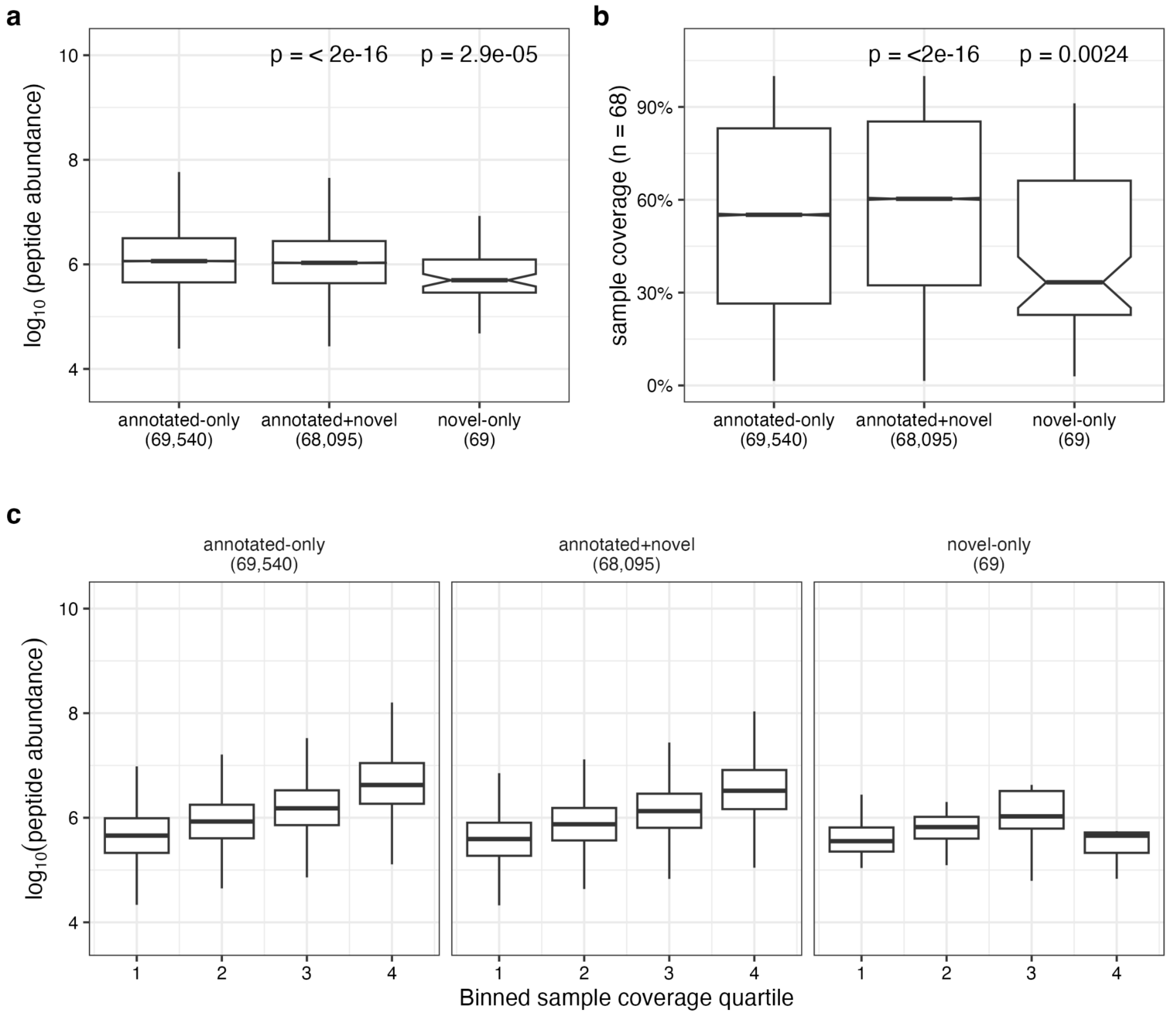


#### Supplementary Figure 5 - Mass spectrometry on human microglia

Peptides split into three groups: those matching to annotated isoforms (annotated-only), matching to both annotated and novel isoforms (annotated+novel) and only to novel isoforms (novel-only). **a**) Comparing the three sets by abundance. **b**) Comparing the three sets by proportional coverage (out of 68 samples). **c**) Binning each set into quartiles based on coverage (x-axis) and visualizing the peptide abundance (y-axis). Box plots show the median and the first and third quartiles of the distribution, with whiskers extending to 1.5 times the interquartile range. Notches denote 95% confidence interval around the median. Peptides falling outside of the whiskers are not plotted. P-values shown are nominal P-values from a Wilcoxon non-parametric test comparing each peptide set to the annotated-only.

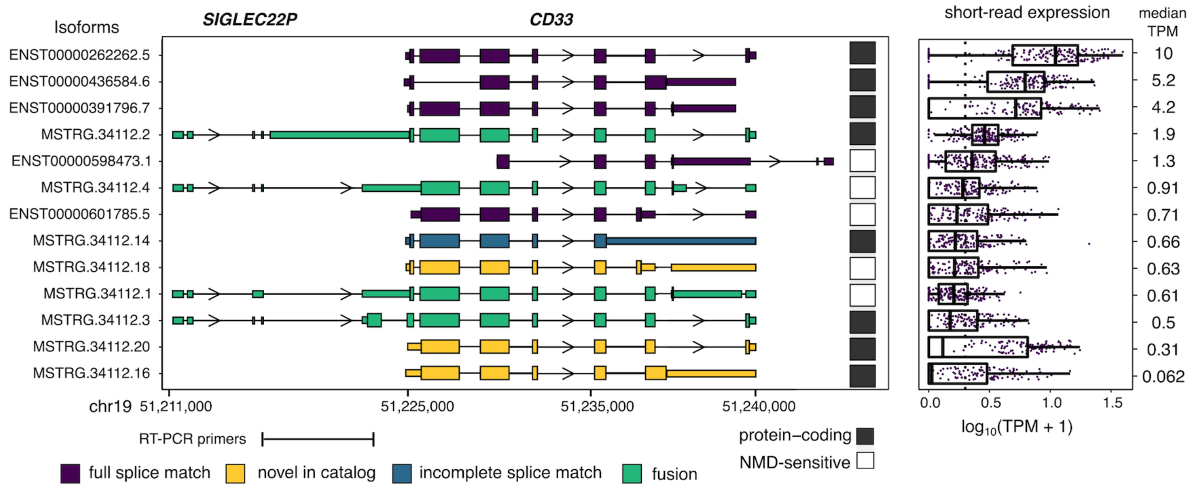


#### Supplementary Figure 6 - CD33 isoforms

All isoforms discovered in long-read RNA-seq mapping to *CD33* and the upstream *SIGLEC22P* gene. Isoform structures are coloured by their SQANTI annotation classification. Short-read expression of each isoform in the single largest microglia cohort (n=185) is plotted on the left, with median expression in transcripts per million highlighted.

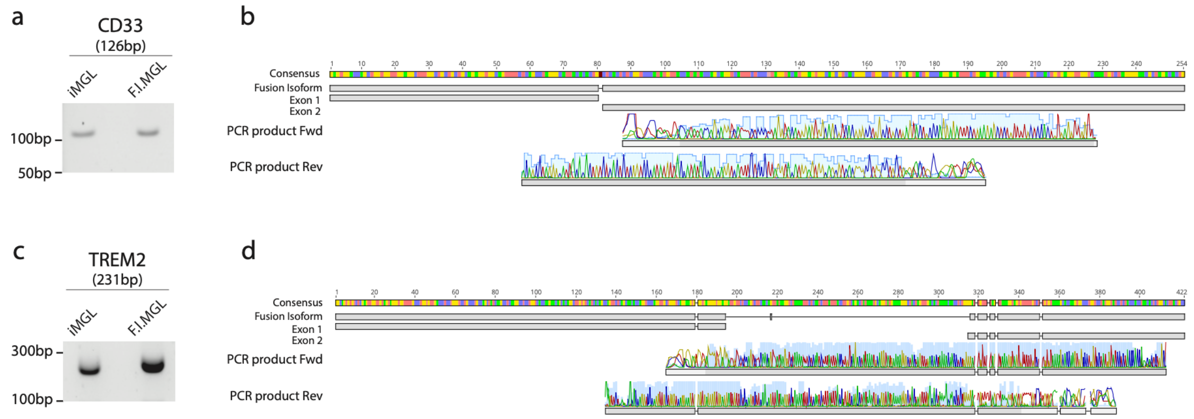


**Supplementary Figure 7 - RT-PCR confirmation of fusion isoforms in *TREM2* and *CD33*.**

a, Bands resulting from the RT-PCR using primers spanning the exon-exon junction for the novel fusion *SIGLEC22P-CD33* isoform in hPSC-derived microglia (iMGL) and freshly isolated microglia (F.I.MGL). b, Sanger sequencing results and alignment with the predicted fusion isoforms. c, PCR band for *TREM2-TREML1* fusion isoform d, Sanger sequencing result and alignment with the predicted isoforms.

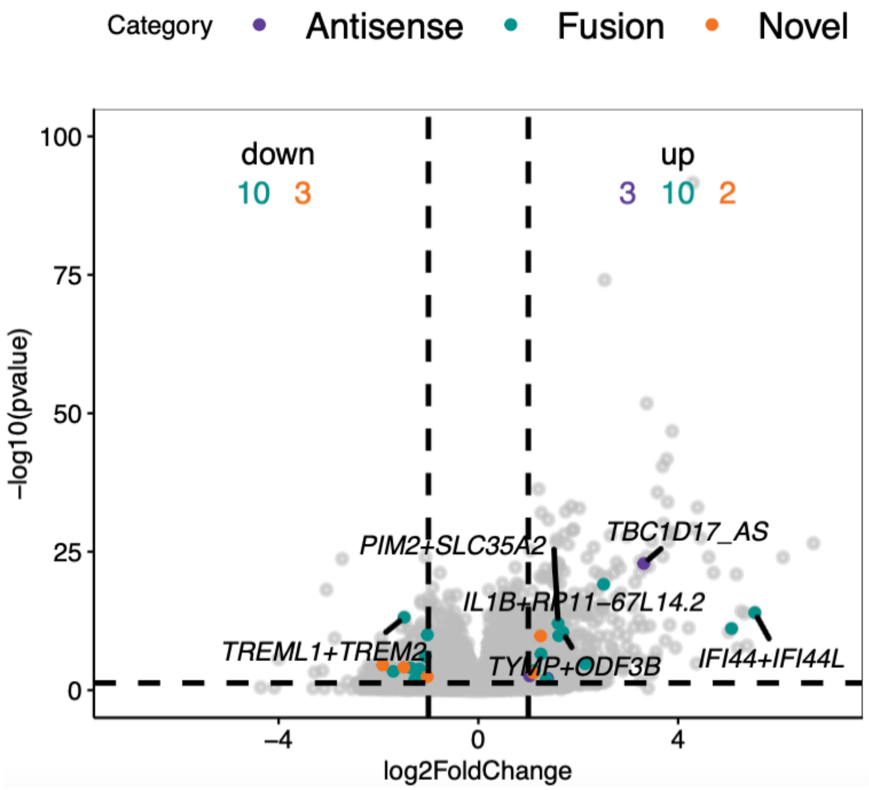


**Supplementary Figure 8 - Differential gene expression in IPS-derived microglia (iMGLs) in response to LPS treatment.**

Volcano plot shows differentially expressed genes at thresholds of |log2 fold change| > 1 and FDR-adjusted P-value < 0.05. Annotated (known) genes are in grey whereas novel genes are coloured.

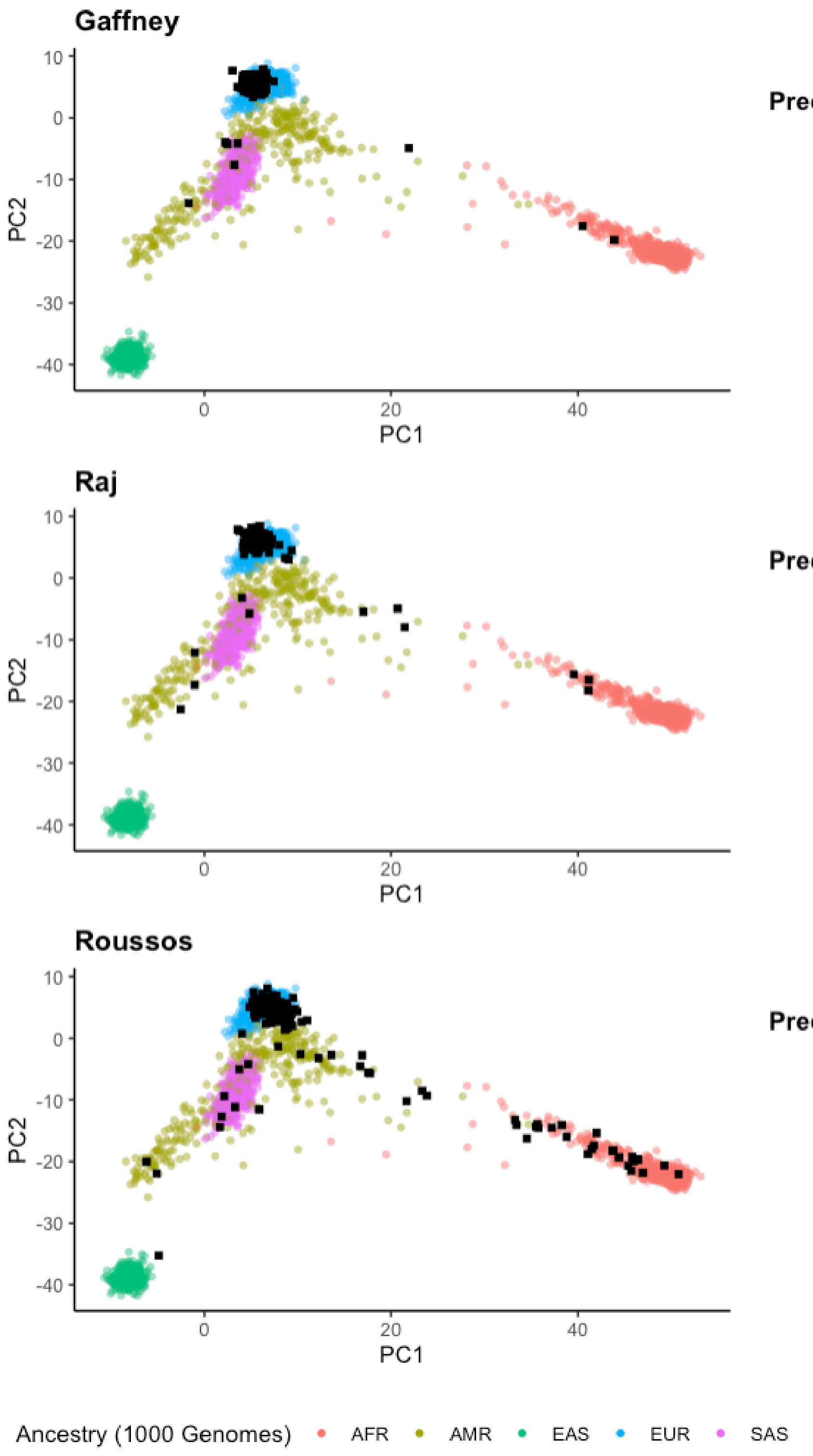


**Gaffney**

| Predicted ancestry | N |
|---|---|
| AFR | 2 |
| AMR | 1 |
| EAS | 1 |
| EUR | 101 |
| SAS | 3 |
| Total | 108 |

**Raj**

| Predicted ancestry | N |
|---|---|
| AFR | 3 |
| AMR | 4 |
| EAS | 3 |
| EUR | 92 |
| SAS | 1 |
| Total | 103 |

**Roussos**

| Predicted ancestry | N |
|---|---|
| AFR | 25 |
| AMR | 16 |
| EAS | 1 |
| EUR | 139 |
| SAS | 4 |
| Total | 185 |

**Supplementary Figure 9 - Predicted ancestries in each cohort.**

All donor genotypes combined together with 1000 genomes reference populations and top two principal components computed. Each point is a donor. Coloured circles are from 1000 genomes reference populations, with black circles the donors from each of the three microglia cohorts. Breakdown of each cohort Ancestry predictions from Somalier. AFR: African, AMR: Admixed American, EAS: East Asian, EUR: European, SAS: South Asian.

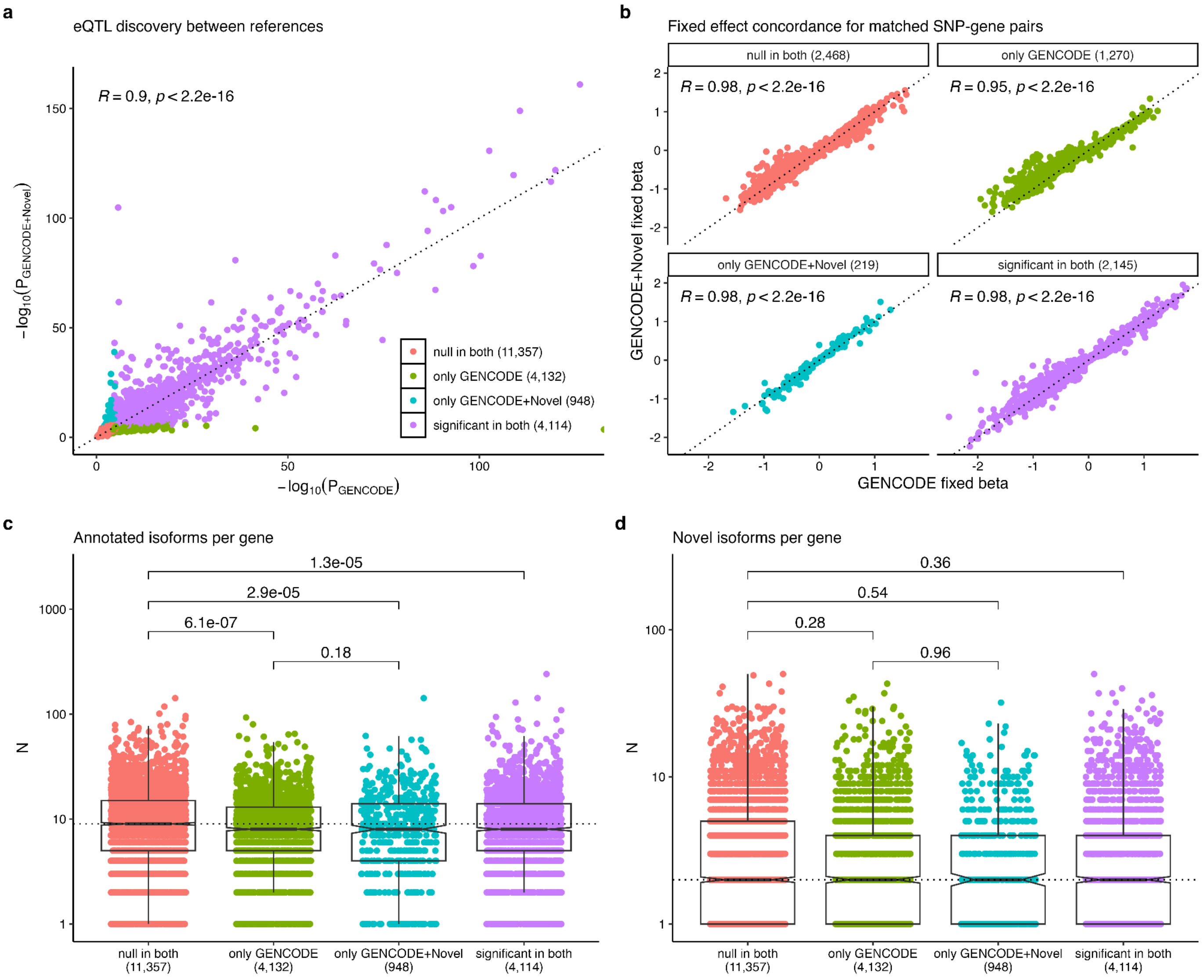


**Supplementary Figure 10 - eQTL discovery between the two references.**

a) Comparing P-values ($-\log_{10}$) for all genes tested in both the GENCODE and GENCODE+Novel reference. Significance threshold was set at Storey's qvalue < 0.05. b) For the SNPs identified as the top cis association in both references, the direction and magnitude of effect is highly concordant, even when the association was not significant in either reference. c) No difference in distribution of numbers of annotated isoforms found between the genes found significant in only one reference. d) No difference seen in distribution of novel isoforms found between the genes found significant to only one reference. P-values from Wilcoxon non-parametric test. Boxplots refer to the first quartile, the median, and third quartile of the distribution. Whiskers extend to 1.5 times the interquartile range.

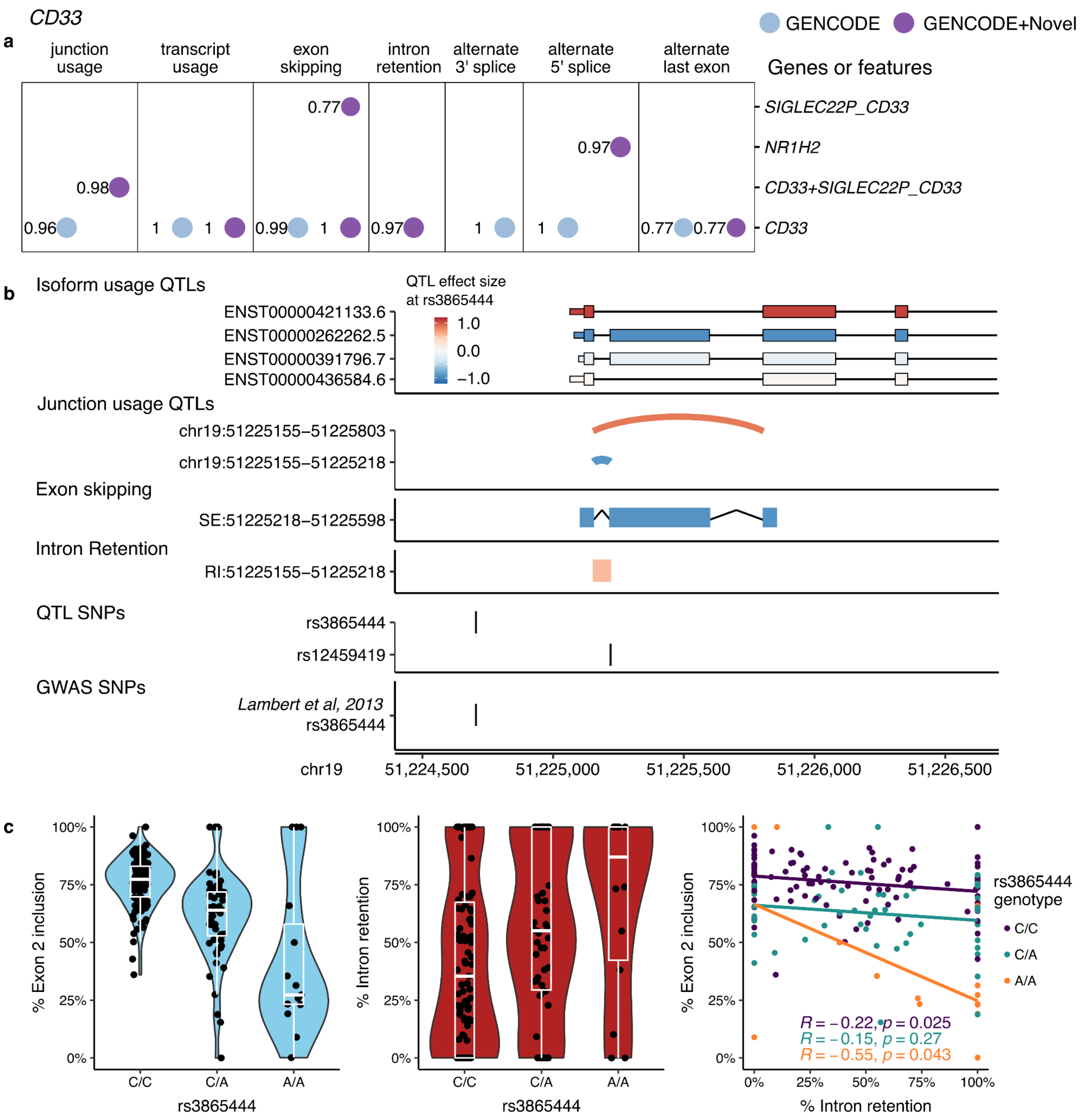


#### Supplementary Figure 11 - splicing QTLs in CD33.

**a**) All sQTL colocalizations at the CD33 locus in the 2 different isoform references. **b**) Colocalized QTLs converge on the skipping of exon 2, as well as increased retention of intron 1. **c)** The lead GWAS SNP associates with reduced exon 2 inclusion and increased intron 1 retention, which are correlated splicing events.

Pedersen, Brent S., Preeti J. Bhetariya, Joe Brown, Gabor Marth, Randy L. Jensen, Mary P. Bronner, Hunter R. Underhill, and Aaron R. Quinlan. n.d. “Somalier: Rapid Relatedness Estimation for Cancer and Germline Studies Using Efficient Genome Sketches.” https://doi.org/10.1101/839944.

Trubetskoy, Vassily, Antonio F. Pardiñas, Ting Qi, Georgia Panagiotaropoulou, Swapnil Awasthi, Tim B. Bigdeli, Julien Bryois, et al. 2022. “Mapping Genomic Loci Implicates Genes and Synaptic Biology in Schizophrenia.” *Nature* 604 (7906): 502–8.

Yao, Douglas W., Luke J. O’Connor, Alkes L. Price, and Alexander Gusev. 2020. “Quantifying Genetic Effects on Disease Mediated by Assayed Gene Expression Levels.” *Nature Genetics* 52 (6): 626–33.

Zou, Yuxin, Peter Carbonetto, Gao Wang, and Matthew Stephens. 2022. “Fine-Mapping from Summary Data with the ‘Sum of Single Effects’ Model.” *PLoS Genetics* 18 (7): e1010299.
